# Intrinsic functional brain connectivity changes following aerobic exercise, computerized cognitive training and their combination in healthy late-middle-aged adults: the Projecte Moviment

**DOI:** 10.1101/2023.05.30.23290731

**Authors:** Stavros I. Dimitriadis, Alba Castells-Sánchez, Francesca Roig-Coll, Rosalía Dacosta-Aguayo, Noemí Lamonja-Vicente, Pere Torán-Monserrat, Alberto García-Molina, Gemma Monte-Rubio, Chelsea Stillman, Alexandre Perera - Lluna, Maria Mataró

**Affiliations:** Department of Clinical Psychology and Psychobiology, University of Barcelona, Barcelona, Spain; Institut de Neurociències, University of Barcelona, Barcelona, Spain; Unitat de Suport a la Recerca Metropolitana Nord, Fundació Institut Universitari per a la recerca a l’Atenció Primària de Salut Jordi Gol i Gurina, Mataró, Spain; Institut d’Investigació en Ciències de la Salut Germans Trias i Pujol (IGTP), Badalona, Spain; Institut de Recerca Sant Joan de Déu, Esplugues de Llobregat, Spain; Department of Medicine, Universitat de Girona, Girona, Spain; Institut Guttmann, Institut Universitari de Neurorehabilitació, Universitat Autònoma de Barcelona, Badalona, Spain; Centre for Comparative Medicine and Bioimage (CMCiB), Germans Trias i Pujol Research Institute (IGTP), Badalona, Spain; Department of Psychology, University of Pittsburgh, Pittsburgh, PA, United States; B2SLab, Departament d’Enginyeria de Sistemes, Automàtica i Informàtica Industrial, Universitat Politècnica de Catalunya, CIBER-BBN, Barcelona 08028, Spain; Department of Biomedical Engineering, Institut de Recerca Pediàtrica Hospital Sant Joan de Déu, Esplugues de Llobregat, Barcelona 08950, Spain; PROFITH “PROmoting FITness and Health Through Physical Activity” Research Group, Department of Physical and Sports Education, Faculty of Sport Sciences, Sport and Health University Research Institute (iMUDS), University of Granada, Granada, Spain; Research Institute, AdventHealth, Orlando, FL, United States

**Keywords:** aerobic exercise, computerized cognitive training, combined training, fMRI, resting-state functional connectivity, default mode network, multiplexity

## Abstract

Lifestyle interventions have positive neuroprotective effects in aging. However, there are still open questions about how changes in resting-state functional connectivity (rsFC) contribute to cognitive improvements. The Projecte Moviment is a 12-weeks randomized controlled trial of a multimodal data acquisition protocol that investigated the effects of aerobic exercise (AE), computerized cognitive training (CCT), and their combination (COMB). An initial list of 109 participants was recruited from which a total of 82 participants (62% female; age = 58.38 ± 5.47) finished the intervention with a level of adherence > 80%. We report intervention-related changes in rsFC, and their potential role as mediators of cognitive benefits. For the AE group, we revealed a limited network of 11 connections showing an increased rsFC that involved mainly the anterior default mode network (aDMN), the posterior default mode network (pDMN : left middle temporal gyrus, and right precuneus), and a decreased rsFC that involved the pDMN (left fusiform gyrus (pDMN), right middle temporal gyrus (pDMN), left and right precentral gyrus), the hippocampus, bilateral supplementary motor areas, and the right thalamus. For the CCT group, we untangled a limited network of 6 connections as a combination of increased and decreased rsFC between brain areas located mainly in the aDMN and pDMN. In the COMB group, we revealed an extended network of 33 connections that involved an increased and decreased rsFC within and between the aDMN/pDMN and a reduced rsFC between the bilateral supplementary motor areas and the right thalamus. No global and especially local rsFC changes due to any intervention mediated the cognitive benefits detected in the AE and COMB groups. Projecte Moviment provides evidence of the clinical relevance of lifestyle interventions and the potential benefits when combining them.

## Introduction

*‘Might strength, efficiency, and segregation update, if you challenge yourself.’* Late adulthood is known for being the finest moment for crystallized cognition; older adults maintain or even increase vocabulary and expertise-based abilities. However, even healthy normal aging, characterized by the lack of pathology, is related to a progressive decline in fluid cognitive abilities, which is associated with reductions in executive function, memory, attention, and processing speed (Buckner, 2004; Craik and Bialystok, 2006; Craik and Byrd, 1982; Glisky, 2007; Harada et al., 2013; Hasher and Zacks, 1988; Kramer et al., 1994; Park et al., 2002; Salthouse and Maurer, 1996; Salthouse, 2010, 1994; Wecker et al., 2005), and deficits in social and independent functioning (Hausman et al., 2020). Concurrently, one of the well-documented hallmarks of aging is the alteration of brain health. Extensive literature focused on the structural changes, such as loss of gray matter volume and less white matter integrity, and their association with age-related cognitive changes (Fan et al., 2019; Ritchie et al., 2015; Yang et al., 2016) and observed decline in brain function.

Functional brain connectivity emerged as a new promising approach based on the fact that cognitive functions are the result of coordinated activity among distant brain areas that work in networks (Sala-Llonch et al., 2015). Functional magnetic resonance imaging (fMRI) allows for examining temporal synchrony derived from a time-series covariance of blood-oxygen-level dependent (BOLD) signals from multiple regions of interest (ROIs) (Bastos and Schoffelen, 2015; Varangis et al., 2019). In particular, resting-state fMRI (rs-fMRI) aims to identify coherences in the spontaneous fluctuations of low-frequency oscillations of the BOLD signal at rest (Biswal et al., 1995) which might be informative of the brain architecture and individual differences (Fox and Raichle, 2007). Studies found that the brain is organized into a number of networks that show high within-network connectivity and lesser long-range connections between networks in order to maximize its efficiency (Bassett and Bullmore, 2006; Bullmore and Sporns, 2009; Madden et al., 2020). Although several networks have been characterized across subjects using rs-fMRI (Damoiseaux et al., 2006) for several aspects (Hausman et al., 2020), the seven most relevant resting-state networks (RSNs) were defined by Yeo et al. (Yeo et al., 2011): the default mode network (DMN), the frontoparietal control network (FPCN), the cingulo-opercular network (CON) or salience network (SN), the dorsal attention network (DAN), the limbic network (LN), the visual network (VN), and the somatomotor network (SMN) (Hausman et al., 2020; Mancho-Fora et al., 2020; Sala-Llonch et al., 2015; Varangis et al., 2019). A general finding is reduced functional connectivity in older adults compared to younger ones (Sala-Llonch et al., 2015) which reflects the reorganization of the brain networks during aging (He et al., 2020). Aging is related to a neural reorganization process characterized by bilateral hemispheric activation of the solicited regions (Hemispheric Asymmetry Reduction in Older Adults model) and decreased functional selectivity and more diffuse and less specialized functional connectivity (Compensation Related Utilization of Neural Circuits Hypothesis model) (Edde et al., 2021; Geerligs et al., 2015; Goh et al., 2012; Joubert and Chainay, 2018; Kang et al., 2021; Madden et al., 2020; Reuter-Lorenz and Lustig, 2005) which has been found to be steeper in clinical populations (Dennis and Thompson, 2014; Farràs-Permanyer et al., 2015). All RSNs experience a certain degree of deterioration in normal aging, but especially the higher-order cognitive networks (Sala-Llonch et al., 2015). Studies found decreased within-network connectivity on the default mode network (DMN), cingulo-opercular network (CON), sensorimotor network (SN), fronto-parietal network (FP) and dorsal-attentional network (DAN), increased connectivity in between networks, less segregated network structure and local efficiency and higher participation coefficient using different types of connectivity analyses (Andrews-Hanna et al., 2007; Bagarinao et al., 2020; Betzel et al., 2014; Chan et al., 2017; Damoiseaux, 2017; Damoiseaux et al., 2008; Geerligs et al., 2015; Grady et al., 2016; Huang et al., 2015; Jones et al., 2011; King et al., 2018; Madden et al., 2020; Onoda et al., 2012; Sala-Llonch et al., 2015; Song et al., 2014; Tomasi and Volkow, 2012; Varangis et al., 2019; Wang et al., 2010). Ongoing research has commonly associated the higher-order cognitive networks (DMN, FPCN, and CON) with deficits in executive function, attention, processing speed, and memory (Andrews-Hanna et al., 2007; Chan et al., 2017; Damoiseaux et al., 2008; Geerligs et al., 2015; King et al., 2018; Onoda et al., 2012; Sala-Llonch et al., 2015; Shaw et al., 2015; Wang et al., 2010). Changes are not that consistent for the somatosensory, motor, and subcortical networks which might remain relatively stable in older adults (Sala-Llonch et al., 2015). Besides, studies involving middle-aged adults found that a decline in connectivity becomes progressively evident (Siman-Tov et al., 2016) with a decrease in segregation (Edde et al., 2020). This fact suggests that middle age might be a critical moment to prevent or delay the inversion of the functional connectivity trajectories and, in turn, promote cognitive health. Results suggest that the last developed areas are the first to be altered, coherent with the hypothesis “last in, last out” (Raz, 2001). Nevertheless, a more integrative approach, the Scaffolding Theory of Aging and Cognition (STAC-r), proposed that multiple factors might influence the age-related decline and an active lifestyle contributes to the scaffolding of novel compensatory networks. This model included behavioral interventions, such as physical and cognitive training, as potential mechanisms to preserve cognitive and brain health and fortunately, emerging evidence indicates that age-related decline may not be inevitable (Stillman et al., 2019).

Different types of exercise have been related to cognitive and brain health. In particular, evidence highlights the role of moderate-intensity aerobic exercise (AE) in the promotion of executive function, processing speed, attention, and memory in older adults (Barha et al., 2017; Colcombe and Kramer, 2003; Etnier and Chang, 2009; Northey et al., 2018). The mechanistic hypotheses of the observed cognitive benefits are multiple and were organized in a three-level model (molecular – brain – behavioral) by Stillman (Stillman et al., 2019) including clear influences of individual variables such as sex and age. At the brain level, while some papers have focused on structural brain changes reporting promising benefits on the volume-specific regions such as the prefrontal cortex and the hippocampus after mid-term interventions (Chaddock-Heyman et al., 2014; Erickson et al., 2014), others have focused on AE-related changes in the functional brain connectivity (Weng et al., 2017). First evidence suggesting an association between physical activity and cardiorespiratory fitness (CRF) with increased coherence in large-scale RSNs, such as DMN, was obtained in cross-sectional samples (Veldsman et al., 2017; Voss et al., 2010a, 2016). Researchers translated this experience to randomized controlled trials (RCT) in order to better understand inconsistencies across different protocols and explore how AE applied as a controlled intervention might produce similar results (Stillman et al., 2019). Although existing RCTs are highly variable and scarce, recent systematic reviews concluded that AE is a viable strategy for modifying functional brain connectivity and perfusion of the hippocampus (Chen et al., 2020; Erickson et al., 2012; Stillman et al., 2019; Tao et al., 2016). Up to now, the most consistent results are found in studies including mid and long-term AE interventions. The longest study, lasting 12 months, found increased efficiency in the DMN, FP, and frontal-executive (FE) networks (Voss et al., 2010b). Six months of AE intervention resulted in increased connectivity between the dorsolateral prefrontal cortex and the superior parietal lobe (Prehn et al., 2019) and a significant association between increased CRF and functional connectivity (FC) between the brain areas of the DMN, although no significant changes were found after the AE in a cross-sectional study (Stillman et al., 2018). Significant results have also been reported after 4 months of interventions showing increased efficiency in the parahippocampus (Tozzi et al., 2016), cerebral blood flow in the hippocampus (Kaiser et al., 2021), and the connectivity between the hippocampus and anterior cingulate cortex (Tao et al., 2019). Shorter AE interventions have shown more inconsistent results. For example, the study of Maass (Maass et al., 2015) found increased perfusion in the hippocampus while Chapman et al. (Chapman et al., 2013) reported non-significant changes in this area but significant in the bilateral anterior cingulate cortex. Three months of cycling have been related to increased rsFC between the DMN and motor regions (McGregor et al., 2018) but 3 months of walking did not lead to significant changes in connectivity between the precuneus and frontal-parietal cortices (Chirles et al., 2017). These results suggest that shorter interventions might be enough to observe changes in brain function although parameters of the physical activity might be better specified. Since all previous 3-month RCTs are scheduled mostly 3 days, we consider that studying changes in brain function in a short-term but high-frequency (5 days) AE intervention might shed light on these inconsistencies.

Cognitive training is another example of the most studied behavioral intervention. Computerized cognitive training (CCT) which refers to single cognitive training tasks performed on electronic devices, became a promising approach to promote cognition and structural and functional brain health (Taya et al., 2015; Ten Brinke et al., 2017). Evidence related CCT to benefits in global cognition as well as specific trained functions such as verbal memory, (Bahar-Fuchs et al., 2017; Barban et al., 2016; Shao et al., 2015), processing speed (Lampit et al., 2014), and executive function (Barban et al., 2016). These results highlighted the potential neuroplasticity even of the aging brain to strengthen or maintain synaptic connections and brain function. Functional brain imaging became a useful technique to describe the neuroplastic mechanisms underlying CCT cognitive effects (Belleville and Bherer, 2012). Most of the papers used a task-fMRI approach, which generally identified decreased activity in the areas functionally involved in the trained task interpreted as more neural efficiency (de Oliveira Rosa et al., 2020; Huntley et al., 2017; Kim et al., 2017). However, literature regarding changes in the spontaneous fluctuations after CCT interventions specifically in healthy older adults is scarce (Cao et al., 2016; Li et al., 2016; Luo et al., 2016). For a detailed review, see van Balkom et al. 2020. Results from previous RCTs found patterns of increased or decreased rsFC involving areas of the DMN, CEN, and DAN after CCT (Chapman et al., 2015; Lampit et al., 2015; Ross et al., 2019; Strenziok et al., 2014). Only one research project including 3 months of multimodal cognitive training applied in the hospital by experts studied the effects of the intervention in multiple age-sensitive networks (Cao et al., 2016). Results showed maintained or increased anterior-posterior and interhemispheric rsFC within the DMN, CEN, and SN maintained DMN-SN coordination and anti-correlation between DMN and CEN (Cao et al., 2016), and more integrated local FC in the training group than controls (Deng et al., 2019). However, to our knowledge, the extent of these effects on home-based computerized multimodal training has not been studied before. We consider that addressing this gap could support CCT as a promising intervention able to counteract age-related rs-FCdecline.

AE and CT showed promising results and a certain degree of complementarity in matters of mechanisms involved in the observed cognitive benefits (Ten Brinke et al., 2019). Exercise impacts most of the body systems promoting low inflammation and oxidative stress, cardiovascular adaptations, and neural repairing responses that might be enhanced by the stimulation and regulation of neuroplasticity through cognitive stimulation (Fabel et al., 2009; Olson et al., 2006). Although evidence is still too few and sometimes inconsistent, systematic reviews argue in favor of an advantage when combining AE and CT (Bamidis et al., 2015; Joubert and Chainay, 2018; Kraft, 2012). Results show that general cognitive function (Shatil, 2013), executive function (Eggenberger et al., 2015), processing speed, and memory (Fabre et al., 2002) benefit from a combined intervention (COMB) and changes in brain structure (Lövdén et al., 2012) and function have been reported.

Six months of combined training have been shown to produce changes in the strength of functional connectivity of the precuneus, right angular gyrus, and posterior cingulate cortex in the DMN and the left frontal eye field in the DAN (Pieramico et al., 2012). Moreover, six months of COMB specifically impacts the connectivity between the medial prefrontal cortex and medial temporal lobe in the DMN (Li et al., 2014). Up to now, just one published paper explored changes in brain function after a shorter period of time but used a task-related fMRI approach. A dual-task training involving AE and working memory training for 12 weeks reported increased brain activity around the bilateral temporoparietal junctions which are highly related to attentional processes, while participants were performing a working memory task in the scanner (Takeuchi et al., 2020). Based on this positive evidence, and the lack of currently published results showing changes in rs-fMRI after short-term COMB interventions, we consider it imperative to analyze potential brain function changes at rest.

Projecte Moviment is an RCT protocol that involved the effect of a high-frequency (5 days per week) short-term (12 weeks) program of AE, computerized cognitive training (CCT), and their combination in healthy physically inactive older adults (Castells-Sánchez et al., 2019). The observed changes in cognition, psychological status, physical activity, molecular biomarkers, and brain volume outcomes have been published in Roig-Coll et al. (Roig-Coll et al., 2020) and Castells-Sánchez et al. (Castells-Sánchez et al., 2022).

In this study, 1) we aim to assess changes in functional connectivity on rs-fMRI related to intervention, 2) we intend to investigate the moderating role of sex and age on functional connectivity on rs-fMRI changes, and 3) the possibility that changes in functional connectivity on rs-fMRI outcomes mediate the relationship between the intervention and cognitive benefits.

## 2. Methods

### 2.1. Study Design

Projecte Moviment is a multi-center, single-blind, proof-of-concept RCT recruiting healthy low active late-middle-aged adults to be assigned in a four parallel-group design including 12-week intervention programs. Participants were assessed at baseline and at trial completion and randomly assigned to an AE group, a CCT group, a COMB group, and a waitlist control group. The study was developed by the University of Barcelona in collaboration with Institut Universitari d’Investigació en Atenció Primària Jordi Gol, Hospital Germans Trias i Pujol and Institut Guttmann, and approved by the responsible ethics committees (Bioethics Commission of the University of Barcelona –IRB00003099- and Clinical Research Ethics Committee of IDIAP Jordi Gol-P16/181-) following the Declaration of Helsinki. The study took place between November 2015 and April 2018.

This research paper follows the previously registered (ClinicalTrials.gov; NCT031123900) and published protocol (Castells-Sánchez et al., 2019). Results on the primary and partial secondary hypotheses (Castells-Sánchez et al., 2021; Roig-Coll et al., 2020) would be included for discussion.

### 2.2. Participants

Healthy adults aged 50 to 70 years old from the Barcelona metropolitan area were recruited using multiple strategies (lists of patients of general physicians, volunteers from previous studies, oral presentations in community centers, advertisements, and local media). Volunteers were informed and screened over the phone and in an on-site interview and those meeting inclusion and exclusion criteria (see Table 1) signed a written informed consent prior to the study involvement.

**Table 1.** Inclusion and exclusion criteria for Projecte Moviment.

| Inclusion criteria | Exclusion criteria |
| --- | --- |
| Aged 50–70 years | Current participation in any cognitive training activity or during last 6 months > 2 h/week |
| ≤120 min/week of physical activity during last 6 months | Diagnostic of dementia or mild cognitive impairment |
| Mini-Mental State Examination (MMSE) $\geq 24$ | Diagnostic of neurological disorder: stroke, epilepsy, multiple sclerosis, traumatic brain injury, brain tumor |
| Montreal Cognitive Assessment 5-min (MoCA 5-min) $\geq 6$ | Diagnostic of psychiatric illness current or during last 5 years |
| Competency in Catalan or Spanish | Geriatric Depression Scale (GDS-15) $> 9$ |
| Adequate visual, auditory, and fine motor skills | Consumption of psychopharmacological drugs current or during last 5 years; or more than 5 years throughout life |
| Acceptance of participation in the study and signature of the informed consent | History of drug abuse or alcoholism current or during last 5 years; or more than 5 years throughout life; $>28$ men and $>18$ woman unit of alcohol/week |
|  | History of chemotherapy |
|  | Contraindication to magnetic resonance imaging |

We randomly assigned participants to AE, CCT, COMB, and waitlist control groups after baseline assessments using a random combination of selected demographic variables (sex, age, and years of education) in order to obtain balanced groups. The allocation sequence was designed by a statistician and the intervention team was responsible for the allocation. Professionals involved in the assessment remained blind to group assignment.

### 2.3. Interventions

Participants assigned to intervention groups went through home-based programs lasting 12 weeks, 5 days per week. Participants randomized to the waitlist control group were on the waitlist for 12 weeks and were asked not to alter their regular lifestyle.

The AE intervention program consisted of a progressive brisk walking program (Week 1: 30 min per day at 9–10 on the Borg Rating of Perceived Exertion Scale (BRPES; (Borg, 1982) perceived as light intensity. Week 2: 45 min per day at 9-10 on BRPES. Week 3 to 12 (10 weeks): 45 min per day at 12-14 on BRPES perceived as moderate-high effort). The CCT intervention program consisted of multimodal cognitive training scheduled in sessions of 45 minutes. Participants used Guttmann Neuropersonal Trainer online platform (GNPT^®^, Spain; Solana et al., 2014, 2015) and performed tasks involving executive function, visual and verbal memory, and sustained, divided, and selective attention. Baseline cognitive performance and the ongoing scores of the activities were used by the GNPT platform to adjust the demand of the activities. The COMB intervention program consisted of a combination of the brisk walking program and the CCT as described above, separately, in single continuous bouts of 45 minutes for each intervention 5 days per week without order or time-point restrictions.

The intervention team was available for participants, registered participants’ activity (phone calls every 2 weeks and a midpoint visit, and a final visit) and ensured participation in solving inconveniences and barriers, and obtained adherence based on participants’ feedback and platform data. Participants monitored their activity in a diary, registering the date and duration of the activity and any adverse events occurring as well as the intensity of the walking in BRPES units.

The protocol for each intervention condition is explained in more detail elsewhere (Castells-Sánchez et al., 2019).

### 2.4. Assessment

Participants went through all assessments in clinical environments at baseline within 2 weeks prior to the start of the intervention and, again, within 2 weeks after the completion of the program (Castells-Sánchez et al., 2019).

#### Neuroimaging: Resting-State Functional MRI data acquisition

##### The Projecte Moviment

MRI data were collected using a 3T Siemens Magnetom Verio Symo MR B17 (Siemens 243 Healthineers, Erlangen, Germany) located at the Hospital Germans Trias i Pujol. Participants were asked to rest with their eyes closed during the scan session, had head motion constrained, and were offered earplugs to reduce the adverse effects of scanner noise. Resting-state functional BOLD imaging scans were obtained with a gradient echo planar imaging sequence (acquisition time: 8:08 min, voxel: 3.1×3.1×3.0mm, TR/TE: 2000/25 ms, flip angle: 78°, slices: 39, thickness: 3 mm, volumes: 240). We also collected T1-weighted multi-planar reformat sequences (acquisition time: 5:26 min, voxel: 0.9×0.9×0.9mm, TR/TE/TI: 1900/2.73/900ms, flip angle: 9°, slices: 192; thickness: 0.9 mm) which were used during the rs-fMRI preprocessing to co-register functional and structural MRI data. Scans were visually checked by an expert neuroradiologist.

##### The NYU Dataset

For evaluating the stability of our findings based on the adopted preprocessing pipeline and functional network construction, we analyzed an open test-retest rs-fMRI dataset. This is an open dataset from the International Neuroimaging Data-Sharing Initiative (INDI) (http://www.nitrc.org/projects/nyu_trt) that was originally described in (Shehzad et al., 2009). The NYU dataset includes 25 participants (mean age 30.7 ± 8.8 years, 16 females) with no history of psychiatric or neurological illness. 3 resting-state scans were acquired from each participant. Scans 2 and 3 were conducted in a single session with 45 min apart, while the scan 1 took place on average 11 months (range 5-16 months) after scans 2 and 3.

Each scan was acquired using a 3T Siemens (Allegra) scanner, and consisted of 197 contiguous EPI functional volumes (TR = 2000 ms; TE = 25 ms; flip angle = 90°; 39 axial slices; field of view (FOV) = 192 × 192 mm2; matrix = 64 × 64; acquisition voxel size = 3 × 3 × 3 mm3). Participants were instructed to remain still with their eyes open during the scan. For spatial normalization and localization, a high-resolution T1-weighted magnetization prepared gradient echo sequence was also obtained (MPRAGE, TR = 2500 ms; TE = 4.35 ms; TI = 900 ms; flip angle = 8°; 176 slices, FOV = 256 mm).

#### Resting-State Functional MRI preprocessing, and denoising

The preprocessing of the rs-fMRI data from both datasets, ‘Projecte Moviment’ and NYU, was conducted following the same standard workflow implemented in the CONN toolbox (http://www.nitrc.org/projects/conn), version 17f (Whitfield-Gabrieli and Nieto-Castanon, 2012). A standard pipeline of preprocessing, involving the steps described below, was applied for consistent functional network topologies (Luppi et al., 2021): removal of the first 5 volumes to allow for steady-state magnetization; functional realignment, motion correction, and spatial normalization to the Montreal Neurological Institute (MNI-152) standard space with 2×2×2mm isotropic resolution. A denoising procedure was driven by applying the anatomical CompCor (aCompCor) method of removing cardiac and motion artifacts,, by regressing out of each individual’s functional data the first 5 principal components corresponding to white matter signal, and the first 5 components corresponding to cerebrospinal fluid signal, as well as six subject-specific realignment parameters (three translations and three rotations) and their first-order temporal derivatives (Behzadi et al., 2007). The subject-specific denoised BOLD signal time series were first linearly detrended, and band-pass filtered between 0.008 and 0.09 Hz to eliminate both, low-frequency drift effects and high-frequency noise. We did not apply spatial smoothing since all analyses were performed on parcellated data. The voxel-based time series within every ROI were averaged to extract a representative time series per ROI. In the present study, the AAL atlas (Tzourio-Mazoyer et al., 2002) with 90 total brain areas (45 ROIs per hemisphere) was considered.

#### Static Functional Connectivity Network Construction: The Multiplex Way

In our study, the construction of a static functional connectivity network (sFCN) incorporates the wavelet decomposition of voxel-based time series and a distance correlation metric to quantify the multiplexity between two brain areas. We performed a wavelet decomposition on every voxel-based time series within every ROI by adopting the maximal overlap discrete wavelet transform (MODWT) (Dimitriadis, 2022). Wavelet coefficients were extracted for the first four wavelet scales for every voxel-based time series which were further averaged to produce 4 frequency-dependent regional time series. Figure 1 illustrates this procedure for a pair of ROIs. Then, we adopted the distance correlation (DC) metric to quantify the multiplexity coupling strength between every pair of ROIs (Székely and Rizzo, 2013). With the DC metric, one can estimate analytically the corresponding p-value of each coupling. This procedure leads to an sFCN of size 90 x 90 per subject and scans in both datasets.

**Figure 1.**
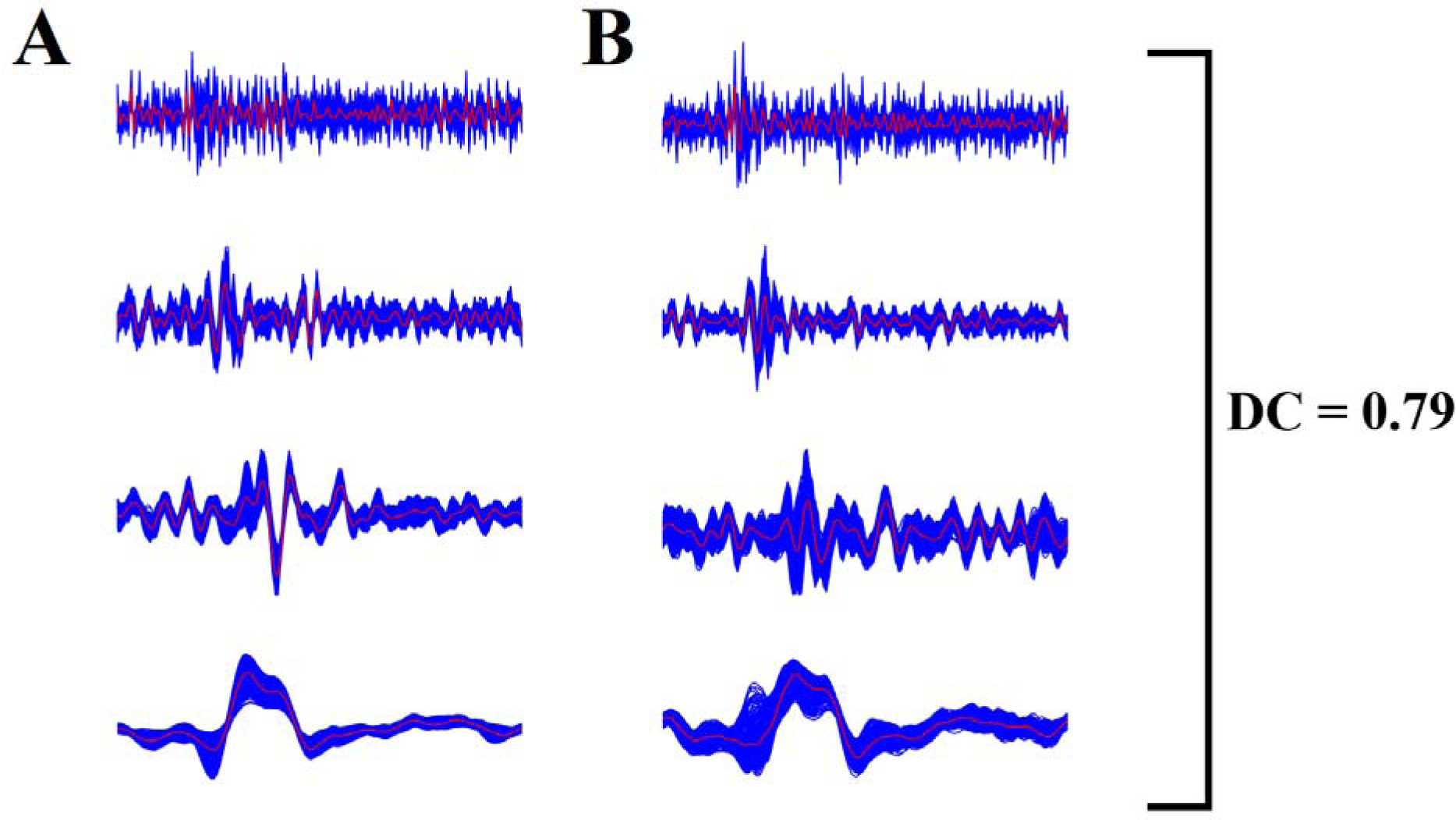
Wavelet decomposition of voxel-based time series and a multiplex coupling strength index. (A,B) A pair set of voxel-based time series (blue) was decomposed with the maximal overlap discrete wavelet transform (MODWT) in four frequency bands. We then averaged the voxel-based time series producing four representative time series per frequency scale and per ROI (red). The distance correlation DC was estimated on the pair of four regional time series.

#### Cognitive Performance

We assessed cognition using a theoretically driven (Lezak et al., 2012; Spreen, 1998) selection of tests that addressed the most relevant cognitive functions: Flexibility (Trail Making Test B-A time; (Tombaugh, 2004)), Fluency (letter and category fluency; (Peña-Casanova et al., 2009)), Inhibition (interference-Stroop Test; (Golden, 2001), Working Memory (backward-WAIS-III; (Wechsler, 2001), Visuospatial Function (copy accuracy-Rey Osterrieth Complex Figure; (Rey, 2009), Language (Boston Naming Test-15; (Goodglass et al., 2001), Attention (forward span, digit symbol coding, and symbol search WAIS-III; (Wechsler, 2001), Speed (Trail Making Test-A; (Tombaugh, 2004); copy time-Rey Osterrieth Complex Figure; (Rey, 2009), Visual Memory (memory accuracy-Rey Osterrieth Complex Figure; (Rey, 2009) and Verbal Memory (total learning and recall-II Rey Auditory Verbal Learning Test; (Schmidt, 1996). Six general domains were designed: 1) Executive Function, 2) Visuospatial Function, 3) Language, 4) Attention-Speed, 5) Memory, and 6) Global Cognitive Function. The cognitive assessment was conducted before the CRF test or any type of exercise to control for acute exercise’s effect on cognitive performance. Extended details in Supplementary Material Table 1.

#### Physical Activity

We obtained physical activity levels with the Minnesota Leisure Time Physical Activity Questionnaire (VREM; (Ruiz Comellas et al., 2012) in which frequency and duration during the last month of multiple activities -sportive walking, sport/dancing, gardening, climbing stairs, shopping, are asked. Then, we calculated energy expenditure for each activity transforming hours per month into units of the metabolic equivalent of tasks (METs). We derived a measure of Sportive Physical Activity (S-PA) by adding METs spent in sportive walking and sport/dancing activities and a measure of Non-Sportive Physical Activity (NS-PA) by summing METs spent in gardening, climbing stairs, shopping, walking, and cleaning house.

#### Cardiorespiratory Fitness

CRF was assessed using the Rockport 1-Mile Test and estimated the maximal aerobic capacity (VO_2max_) using the standard equation reported by Kline et al. (Kline et al., 1987). The equation uses the following variables to estimate VO_2max_: weight, age, sex, time to complete a mile, and heart rate at the end of the test. During the test, participants were instructed to walk one mile on a treadmill adjusting their speed in order to be as fast as possible without running.

### 2.5. Statistical Analysis

Statistical procedures were performed using IBM SPSS Statistics 27. The distribution of raw scores was assessed to ensure data quality (i.e., outliers, skewness). We calculated change scores (post-test minus pre-test), and compared baseline scores between groups.

#### Detecting Functional Connectivity Differences between Pre and Post-Intervention Time Periods

1) We applied ANOVA between baseline global mean strength of sFCN for the four groups (AE, CCT, COMB, and control) to assess potential baseline differences; We also applied ANOVA between baseline and follow-up global mean strength of sFCN for the four groups. We adopted a similar approach between every pair of scans for the test-retest NYU dataset.
2) To reveal functional connectivity differences due to intervention, we applied a Wilcoxon Rank-Sum Test between pre and post-intervention related sFCN independently for every group and for every pair of ROIs. Finally, we corrected for multiple comparisons with a false discovery rate (FDR) with a significance level of p < 0.05 (Benjamini and Hochberg, 1995). We followed a similar approach between every pair of scans for the test-retest NYU dataset.
3) sFCN difference maps (post-test minus pre-test) were compared between the intervention group and the control (AE vs control; CCT vs control; COMB vs control) by adopting a Wilcoxon rank-sum test; Results were corrected for multiple comparisons using FDR, with a significant level of p < 0.05.

#### Assessing the Repeatability of sFCN from the NYU Dataset

4) We applied similar statistical analyses as described in 1) and 2) between the scan-based sFCN for the NYU dataset to assess the repeatability of the functional connectivity patterns as derived from the adopted pipeline.

#### Moderating and Mediation Analysis

5) We adopted the PROCESS Macro for SPSS (Hayes and Rockwood, 2016) to analyze the moderating effect of age and sex on intervention-related changes for global mean DC strength estimated over the individual sFCN.
6) We also used the PROCESS macro to perform mediation analyses to assess whether a change in the global mean DC strength estimated over the individual sFCN mediated the cognitive benefits observed in the AE and COMB groups (Roig-Coll et al., 2020). These benefits include for the AE group, the Executive Function (Working Memory) and Attention-Speed (Attention) and in the COMB group, the changes in Attention-Speed (Attention and Speed).
7) Complementary, our mediation analysis was also driven by the outcome of the analysis over the sFCN difference maps (post-test minus pre-test) where the comparison between pre-post periods for each active group (AE-pre vs AE-post; CCT-pre vs CCT-post; COMB-pre vs COMB-post) revealed a subnetwork of functional connections that increased (positive network) or decreased (negative) due to the intervention. The mean DC strength of either the positive or negative subnetwork acted as mediators.
8) We also explored if the outcome of the analysis over the sFCN difference maps (post-test minus pre-test) mediated the relationship between the changes in the CRF and changes in the cognitive measures targeting on the Executive Function (Working Memory) and the Attention-Speed (Attention).

For these mediation analyses, we created a treatment variable (condition vs control) as the independent variable, change in cognition for those functions that showed significant intervention-related changes as the dependent variable, and changes in the CRF, the global mean DC strengths and the mean DC strength of the positive/negative subnetwork have been the mediators controlling for baseline performance score, age, sex and years of education. These analyses were computed with bias-corrected bootstrapped 95% confidence intervals (CIs) based on 5,000 bootstrap samples. The significance was indicated if the CIs in Path AB did not overlap with 0 (Hayes and Rockwood, 2016).

## 3. Results

### 3.1 Participants

A total of 109 participants completed the baseline assessment and 92 completed the intervention (intention to treat sample, ITT) (see Figure 1 in (Roig-Coll et al., 2020)). The Per Protocol (PP) sample, analyzed in the present study, included 82 subjects (62% female; age = 58.38 ± 5.47) with a level of adherence > 80%. The demographics of the PP sample are tabulated in Table 2. There were no participants’ differences at the baseline across the groups in physical and cognitive outcomes except for Non Sportive Physical Activity (NS-PA) and current smoking status (see supp. material Tables 2.1, 2.2 for extended details). Current smoking status at baseline was included as a covariate.

**Table 2.** Participants Characteristics at Baseline

|  | **Total**<br>Mean (SD) | **AE**<br>Mean (SD) | **CCT**<br>Mean (SD) | **COMB**<br>Mean (SD) | **Control**<br>Mean (SD) | **Group**<br>**Comparison** |
| --- | --- | --- | --- | --- | --- | --- |
| n total / n females | 82 / 51 | 25 / 13 | 23 / 16 | 19 / 14 | 15 / 8 | $\chi^2(3) = 3.20$,<br>$p = .361$ |
| Age (years) | 58.38<br>(5.47) | 58.40<br>(5.12) | 57.91<br>(5.31) | 60.32 (5.54) | 56.60 (5.97) | $H(3) = 3.53$,<br>$p = .317$ |
| Years of education | 12.52<br>(5.57) | 12.44<br>(5.75) | 12.04<br>(4.94) | 12.37 (5.43) | 13.60 (6.72) | $H(3) = 0.28$,<br>$p = .963$ |

AE = Aerobic Exercise; BMI = Body Mass Index; CCT = Computerized Cognitive Training; COMB = Combined Training; WAIS-III = Wechsler Adult Intelligence Scale. Mean (SD); $\chi^2$ = chi square; H = Kruskal Wallis H test; F = Anova test. See Supplementary Tables 4 for more cognitive, physical, cardiovascular risk factors, global FA and MD outcomes at Baseline.

### 3.2 Repeatable sFCN Produced by the Adopted Pipeline

Our analytic pipeline revealed consistent mean functional connectivity coupling strength between short-term (mins) and long-term (months) scans as estimated over the rs-fMRI test-retest NYU dataset (see supp. Table 2.3). Moreover, we didn’t detect any significant difference between every pair of scans on the ROI-ROI level. Both analyses, on the global and local levels, favor the adaptation of the pipeline for the analysis of the rs-fMRI recordings from our intervention study.

### 3.3 Intervention-related Changes in static Functional Connectivity

The global mean strength of sFCN did not differ significantly between the four groups at the baseline (see supp. material Table 2.3). Moreover, the global mean strength of the sFCN for the three groups (AE, CCT, and control) did not differ significantly between the baseline and the follow-up but it differs only for the COMB group (see supp. material Table 3).

Regarding the pre-post group analysis on the ROI level, Figure 2 illustrates the significant connections that survived the statistical thresholds for every intervention group comparing the baseline and the follow-up conditions.The AE and CCT interventions led to a small number of connections, 11 and 6 connections, correspondingly while the COMB intervention protocol produced an extended network of 33 connections (Fig.2). Our findings involved a combination of increased coupling DC strength (red) and decreased coupling DC strength (blue) due to the intervention protocol. The averaged increment and decrement of DC strength over the detected pairs of connections are tabulated in Table 3 showing no group difference.

**Figure 2.**
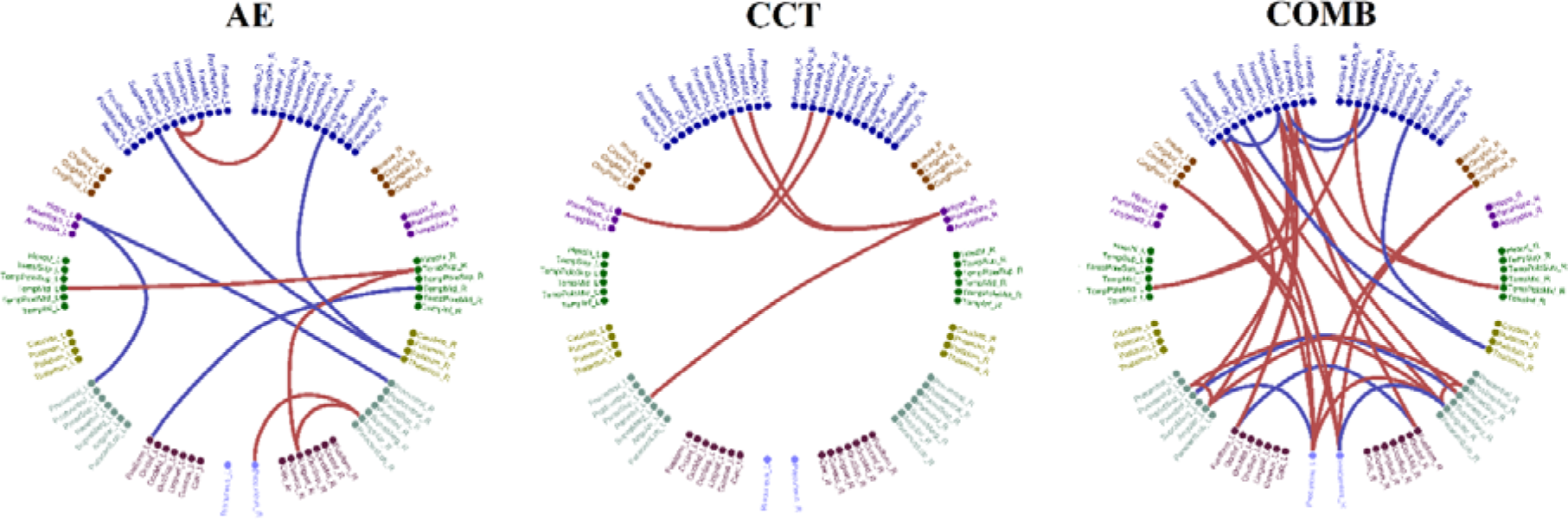
Topological layouts of increased (red) and decreased (blue) rsFC between pairs of ROIs in the AAL space for every intervention group comparing the baseline and the follow-up conditions. (red = increased coupling DC strength due to the intervention blue = decreased coupling DC strength due to the intervention)

**Table 3.** Averaged increment and decrement of DC strength over the detected pairs of connections demonstrated in the Figure 2.

|  | **AE**<br>M(SD) Increased -<br>M(SD) Decreased | **CCT**<br>M(SD) Increased -<br>M(SD) Decreased | **COMB**<br>M(SD) Increased -<br>M(SD) Decreased |
| --- | --- | --- | --- |
| Mean DC Strength (MOVIMENT) | 0.035 (0.005) – -<br>0.021 (0.007) | 0.031 (0.006) | 0.041 (0.006) – - 0.032<br>(0.005) |

AE = Aerobic Exercise; CCT = Computerized Cognitive Training; COMB = Combined Training; M = Mean ; SD = Standard Deviation

Specifically, for the AE group, we revealed an increased rsFC between the right lingual gyrus (Visual Network - VN) and right superior temporal gyrus (ventral attentional network - VAN), and right angular gyrus (fronto-parietal network - FPN), right superior temporal gyrus (ventral attentional network - VAN), and left middle temporal gyrus (posterior DMN - pDMN), right angular gyrus (VN) and right precuneus (pDMN), left and right inferior frontal gyri (orbital part) with left frontal middle gyrus (orbital part) (anterior DMN - aDMN). We also revealed a decreased rsFC between the left fusiform gyrus (pDMN) and right middle temporal gyrus (pDMN), left hippocampus and left and right precentral gyrus (pDMN), and between left and right supplementary motor areas and right thalamus.

For the CCT group, we untangled an increased rsFC between the left hippocampus (pDMN) and right inferior frontal gyrus (opercular part) (aDMN) and right middle frontal gyrus (aDMN), right hippocampus (pDMN) and left middle frontal gyrus (aDMN), and left inferior frontal gyrus (opercular part) (aDMN), left superior frontal gyrus (aDMN) and left inferior parietal gyrus (pDMN).

The strongest changes were observed in the COMB group in brain areas that involved an increased rsFC of within FPN coupling strength between the left and right middle frontal gyri and left and right temporal poles (middle temporal gyri), the increment between left and right superior parietal lobules and left and right angular gyri (FPN), an increased rsFC between left and right middle frontal gyri (FPN) and left and right temporal poles (middle temporal gyri), the increment between posterior DMN and FPN that involved the left/right precuneus (pDMN) with the right angular gyrus (FPN) and with the posterior cingulate gyrus (pDMN), and an increased rsFC between the left portion of the aDMN that involves the left middle frontal gyrus (orbital part), the inferior frontal gyrus (opercular part), the superior frontal gyrus (medial part), and the middle frontal gyrus (orbital part) with the pDMN that involves left/right fusiform gyrus, left/right parietal inferior gyri, left/right angular gyrus, and left/right precuneus. The COMB intervention group showed a decreased rsFC between areas within the pDMN (left and right fusiform gyri, left and right inferior parietal gyri, left angular gyrus - left precuneus, right angular-right precuneus), within the aDMN (left and right middle frontal gyri (orbital part), left and right inferior frontal gyri - opercular part, left inferior frontal gyrus (opercular part) with left superior frontal gyrus (medial part), left superior frontal gyrus (medial part) with left middle frontal gyrus (orbital part) and between the left and right supplementary motor and right thalamus.

The mean and total DC coupling strength of the detected connections within aDMN (decrement), within pDMN (decrement), and between aDMN - pDMN (increment) brain areas will also feed the mediation analysis (see section 3.6).

Statistical analysis of sFCN difference maps (post-test minus pre-test) in a pairwise ROI fashion between the intervention group and the control (AE vs control; CCT vs control; COMB vs control) didn’t reveal any findings.

### 3.4 Sex and age moderation effects

Moderation analyses showed that age did not significantly moderate the effects of the intervention on global mean rsFC strength estimated over the individual sFCN in any group. Sex did not significantly moderate the effects of the intervention on global mean rsFC strength estimated over the individual sFCN in any group.

### 3.5 Mediation effects on intervention-related cognitive benefits

We applied mediation analyses to investigate whether changes in rsFC mediated the association between the intervention and the cognitive domains that demonstrated a significant change as reported in Roig-Coll et al. (Roig-Coll et al., 2020). As rs-FC, we employed the global mean rsFC strengths, the mean, and the total rsFC strength of the positive/negative subnetworks that involved the within aDMN, the within pDMN, and their interactions (aDMN-pDMN). In our previous study, we showed that the AE group showed improvement in Executive Function (Working Memory) and Attention-Speed (Attention) and the COMB group showed changes in Attention-Speed (Attention and Speed). Mediation analyses showed that changes in global mean DC strengths and mean and total DC strength of the positive/negative subnetwork did not significantly mediate the observed cognitive benefits for any group. Finally, the integrated increment and decrement of rsFC changes detected (Figure 2) did not mediate the relationship between CRF and executive Function (Working Memory) and Attention-Speed (Attention).

## 4. Discussion

In this paper, we report intervention-related changes in rs-fMRI static functional connectivity in our Projecte Moviment trial, which investigates the potential neuroprotective effects of interventions (AE, CCT, COMB) in healthy late-middle-aged adults compared to a healthy control group (Castells-Sánchez et al., 2019). In our previous study, we reported cognitive changes in Executive Function and Attention-Speed in the AE group, and Attention-Speed in the COMB group (Roig-Coll et al., 2020).

In our study, the participants who followed the AE program showed positive changes in rs-fMRI functional connectivity in agreement with previous studies (Stillman et al., 2019, 2016). In the present study, we reported an increased rsFC that involved mainly the aDMN, pDMN, and also FPN, VAN, and VN, and negative rsFC changes that involved the pDMN, the hippocampus, and left and right supplementary motor areas and right thalamus. Previous studies reported an association between AE and increased coupling within the DMN (Chirles et al., 2017; McGregor et al., 2018), in the hippocampal network (Won et al., 2021a) and also a decreased rsFC within the DMN (Chirles et al., 2017; McFadden et al., 2013), and the SMN (Flodin et al., 2017). Although the interpretation of the reductions of decreased rsFC is challenging, AE-related decreased rsFC co-occurred with decreased fat mass (McFadden et al., 2013), and cognitive stability (Chirles et al., 2017). It is evident from our findings that longer interventions are needed to induce extended changes in both metabolic, and rs-fMRI connectivity networks. Parameters of the AE are critical for the neuroprotective effects of the exercise which can be altered by sex, age, and health status (Stillman et al., 2020; Won et al., 2021a).

For the CCT group, we reported an increased rsFC between the left hippocampus (pDMN) and right inferior frontal gyrus (opercular part) (aDMN) and right middle frontal gyrus (aDMN), right hippocampus (pDMN) and left middle frontal gyrus (aDMN), and left inferior frontal gyrus (opercular part) (aDMN), left superior frontal gyrus (aDMN), and left inferior parietal gyrus (pDMN). Previous studies in healthy older adults reported a combination of increased rsFC and decreased rsFC in the DMN as a consequence of a CCT program (for a review see (Ten Brinke et al., 2017)). The maintenance of global mean rsFC on the same level in conjunction with an increased rsFC pattern that involved DMN and also the hippocampus could be characterized as a positive outcome of CCT intervention. These observations could be linked to the CCT protocol but also to the general motivation of the subjects that participated in a lifestyle behavior project. In our previous study, (Roig-Coll et al., 2020), participants in the CCT did not show any significant changes in physical activity status, sleep patterns, and psychological health.

Our study also evaluates the benefits of combining multimodal CCT and brisk walking for 12 weeks, 5 days per week in bouts of 54 min. Participants of the COMB group showed an improvement in both global and local rsFC compared to the control group. Our findings supported both the benefits of COMB intervention in terms of both local and global rsFC. The increment of global rsFC is a positive outcome of the COMB intervention by comparing the global mean of our group of old adults compared to the global mean of younger adults from the NYU dataset (see supp. material Table 2.3 for NYU dataset vs supp. material Table 3). Our pre-post analysis of rsFC in the COMB group revealed an extended network of functional connections where their strength either increased or decreased in the follow-up compared to the baseline. This network involved mainly the DMN and to a lesser extent the FPN, and also the supplementary motor area with the thalamus as it was observed in the AE group. Specifically, we observed a decreased rsFC pattern for brain areas located within the aDMN and within the pDMN, and an increased rsFC pattern between brain areas located over the aDMN and pDMN. A reduced rsFC was also detected between the left and right supplementary motor areas and the right thalamus. Our rsFC findings supported the positive outcome of the COMB intervention protocol within the 12 weeks period compared to the rsFC findings in CCT and AE groups. There is no rs-fMRI intervention study that followed the same protocol with three active and a control group in order to directly compare our findings. However, rsFC within the DMN is the most common resting-state network under investigation that has been also found to be impacted by aging (Geerligs et al., 2015; Mevel et al., 2011). In normal aging, various DMN areas like the superior and middle frontal gyrus, superior parietal cortex, and posterior cingulate cortex showed a decreased rsFC (Hafkemeijer et al., 2012). Age-associated changes in rsFC were also observed between the anterior versus the posterior DMN (Andrews-Hanna et al., 2007). In the aDMN, both increment and decrement were revealed in the frontal lobe, whereas within the pDMN only a decreased rsFC pattern was detected associated with aging (Wang et al., 2006) (Wang et al., 2006). It is hypothesized that the increased rsFC in the aDMN could serve as a compensatory mechanism that attempts to balance the loss of cognitive functions (Grady et al., 2003; Wang et al., 2006)). It is the first time in the literature that a study reports changes in rsFC within aDMN, pDMN, and also between aDMN - pDMN brain areas.

In moderation analyses, we did not find a significant moderation effect of age on intervention-related changes in the global mean rsFC strength estimated over the individual sFCN in any group. These findings could be supported by the tight age-range of our sample and the fact that our sample was young compared to targeted clinical populations where the brain health of older subjects tends to decline faster compared to the intervention group (Erickson et al., 2014). We didn’t find a significant moderation effect of sex in the global mean rsFC strength estimated over the individual sFCN in any group. However, more studies from our group and other research groups suggested that biological age acts as a moderator of the relationship between aerobic exercise and neuroprotective effects (Barha and Liu-Ambrose, 2018; Barha et al., 2019; Castells-Sánchez et al., 2022, 2021, 2019). Sex differences in the response of cardiovascular, musculoskeletal, and respiratory systems in AE and the impact of sex hormones may explain the different intervention-related changes in cardiovascular risk factors (Barha et al., 2019; Barha and Liu-Ambrose, 2018).

To reveal the possible mechanisms that could explain the cognitive benefits in the AE group in the Executive Function (Working Memory) and Attention-Speed (Attention) and in the COMB group in the Attention-Speed (Attention and Speed) (Roig-Coll et al., 2020), we explored the mediation effects of the global mean rsFC strength, and the local rsFC findings in the COMB group (the mean rsFC strength within aDMN, within pDMN, and between aDMN - pDMN). No global and especially local rsFC changes due to any intervention mediated the cognitive benefits detected in the AE and COMB groups. Additionally, the integrated increment and decrement rsFC changes due to the intervention protocol (Figure 2) didn’t mediate the relationship between CRF and the executive Function (Working Memory) and Attention-Speed (Attention) measurements.

The failure of the mediation analysis with rsFC as actual mediators could be explained due to the short duration of the intervention and/or the sample size which might not be enough to reveal such mediation effects in the cognitive functions of high variability (Stillman et al., 2016).

### Methodological Considerations

While there is partial consistency in neuropsychological, and structural neuroimaging findings across the studies, results based on rsFC analysis are less consistent. The reason for this inconsistency is due to a large repertoire of available analytic paths that one researcher can follow. Below, we summarize the available rsFC options: the seed-based analysis targeting one or a few seed ROIs (Chirles et al., 2017; Flodin et al., 2017; McGregor et al., 2018; Prehn et al., 2019; Won et al., 2021a, 2021b), the ROI-ROI based on the parcellated space by adopting an atlas (Voss et al., 2010a), the BOLD fluctuation (Tozzi et al., 2016), the graph theory methods (Burdette et al., 2010; Tozzi et al., 2016), and the application of ICA to detect the seven resting-state networks where this approach is called network-of-interest (NOI) to network-of-interest (NOI) (McFadden et al., 2013). The diversity of methodological analytic approaches across those reports is due to an explosion of the proposed alternative methods that one researcher can follow to investigate the rsFC. For that reason, differences between the analytic methods adopted by intervention studies should be taken into consideration in the final interpretation of the converging findings. Specifically, in our study which is the first rs-fMRI study that includes three active groups and a control group, our findings should be replicated by another study following a similar analytic approach. However, to guarantee that our findings are not spurious due to the adopted analytic plan that includes specific network construction steps (Luppi et al., 2021), we ran the same analysis in a test-retest rs-fMRI study with three scans. The consistency of rsFC between scans in the NYU dataset further supports our analytic pipeline and the findings in the Moviment Projecte trial.

Future intervention research should invite a larger number of participants followed by age and sex-balanced groups that will support intra-group comparative analyses. It would be interesting to also register diet patterns that influence cognitive functions and rsFC (Dimitriadis et al., 2021) and can potentially positively affect cardiovascular risk factors, and anthropometric and blood sample measures. Additionally, it would be interesting to register the participants longitudinally at more than two-time points. Our Projecte Moviment will advance our understanding of the intervention protocols by taking the advantage of new omics technology that will further shed light on biological pathways supporting the cognitive benefits as a consequence of the intervention protocol.

In summary, the present study demonstrates the potential benefits of lifestyle interventions when a combined physical and cognitive intervention protocol is adopted. For the AE group, we reported an increased rsFC that involved mainly the aDMN, pDMN, and also FPN, VAN, and VN, and a decreased rsFC that involved the pDMN, the hippocampus, and left and right supplementary motor areas, and the right thalamus. For the CCT group, we found a combination of increased and decreased rsFC between brain areas located mainly in the aDMN and pDMN. The greatest alterations of rsFC were revealed in the COMB group. These findings involved a decreased rsFC within the aDMN, within the pDMN, an increased rsFC between the aDMN - pDMN, and a reduced rsFC between the left and right supplementary motor areas and the right thalamus as it was found in the AE group.

## Data Availability

All data produced in the present study are available upon reasonable request to the PI (Professor Mataro)

## Acknowledgments

SID is supported by a Beatriu de Pinós fellowship (2020 BP 00116). This work was supported by the Spanish Ministry of Economy and Competitiveness (www.mineco.gob.es) PID2021-122952OB-I00, Networking Biomedical Research Centre in the subject area of Bioengineering, Biomaterials and Nanomedicine (CIBER-BBN), initiatives of Instituto de Investigación Carlos III (ISCIII), and Share4Rare project (Grant Agreement 780262). We would like to thank the agreement with Technogym to use their treadmill and Gràfiques Llopis, S.A., for their support on the image design of the project.

## Ethics Statement

This study was carried out in accordance with the recommendations of SPIRIT Guidelines with written informed consent from all subjects. All subjects gave written informed consent in accordance with the Declaration of Helsinki. The protocol was approved by the Bioethics Commission of the University of Barcelona (IRB00003099) and Clinical Research Ethics Committee of IDIAP Jordi Gol (P16/181).

## CRediT authorship contribution statement

MM conceptualized the study and contributed to the study design and implementation as Principal Investigator. SID designed the methodology, the software, the visualization and the writing of the original draft. PT-M made substantial contributions to the design and content of the trial. AC-S, FR-C, and NL-V contributed to the design, implementation, and writing of the protocol. RD-A contributed to the design of the trial from their area of expertise. AG-M and GM-R collaborated in the implementation of specific procedures. All authors reviewed the manuscript and provided the final approval for the manuscript.

## Conflict of Interest Statement

The authors declare that the research was conducted in the absence of any commercial or financial relationships that could be construed as a potential conflict of interest.

## Funding

Projecte Moviment is a project funded by the Spanish Ministry of Economy and Competitiveness under two grants: Neuroplasticity in the adulthood: physical exercise and cognitive training (PSI2013-47724-P) and Integrative omics study on the neurobiological effects of physical activity and cognitive stimulation (PSI2016-77475-R). It has also been rewarded with three pre-doctoral fellowships (FPU014/01460, FI-2016, and FI-2018).

## Notes

### Competing Interest Statement

The authors have declared no competing interest.

### Clinical Trial

NCT031123900

### Clinical Protocols

https://www.frontiersin.org/articles/10.3389/fnagi.2019.00216/full

### Author Declarations

(Bioethics Commission of the University of Barcelona IRB00003099 and Clinical Research Ethics Committee of IDIAP Jordi Gol P16/181) following the Declaration of Helsinki. The study took place between November 2015 and April 2018.

## Bibliography

Andrews-Hanna, J.R., Snyder, A.Z., Vincent, J.L., Lustig, C., Head, D., Raichle, M.E., Buckner, R.L., 2007. Disruption of large-scale brain systems in advanced aging. Neuron 56, 924–935. doi:10.1016/j.neuron.2007.10.038

Bagarinao, E., Watanabe, H., Maesawa, S., Mori, D., Hara, K., Kawabata, K., Yoneyama, N., Ohdake, R., Imai, K., Masuda, M., Yokoi, T., Ogura, A., Taoka, T., Koyama, S., Tanabe, H.C., Katsuno, M., Wakabayashi, T., Kuzuya, M., Hoshiyama, M., Isoda, H., Sobue, G., 2020. Aging impacts the overall connectivity strength of regions critical for information transfer among brain networks. Front. Aging Neurosci. 12, 592469. doi:10.3389/fnagi.2020.592469

Bahar-Fuchs, A., Webb, S., Bartsch, L., Clare, L., Rebok, G., Cherbuin, N., Anstey, K.J., 2017. Tailored and adaptive computerized cognitive training in older adults at risk for dementia: A randomized controlled trial. J Alzheimers Dis 60, 889–911. doi:10.3233/JAD-170404

Bamidis, P.D., Fissler, P., Papageorgiou, S.G., Zilidou, V., Konstantinidis, E.I., Billis, A.S., Romanopoulou, E., Karagianni, M., Beratis, I., Tsapanou, A., Tsilikopoulou, G., Grigoriadou, E., Ladas, A., Kyrillidou, A., Tsolaki, A., Frantzidis, C., Sidiropoulos, E., Siountas, A., Matsi, S., Papatriantafyllou, J., Kolassa, I.-T., 2015. Gains in cognition through combined cognitive and physical training: the role of training dosage and severity of neurocognitive disorder. Front. Aging Neurosci. 7, 152. doi:10.3389/fnagi.2015.00152

Barban, F., Annicchiarico, R., Pantelopoulos, S., Federici, A., Perri, R., Fadda, L., Carlesimo, G.A., Ricci, C., Giuli, S., Scalici, F., Turchetta, C.S., Adriano, F., Lombardi, M.G., Zaccarelli, C., Cirillo, G., Passuti, S., Mattarelli, P., Lymperopoulou, O., Sakka, P., Ntanasi, E., Caltagirone, C., 2016. Protecting cognition from aging and Alzheimer’s disease: a computerized cognitive training combined with reminiscence therapy. Int. J. Geriatr. Psychiatry 31, 340–348. doi:10.1002/gps.4328

Barha, C.K., Davis, J.C., Falck, R.S., Nagamatsu, L.S., Liu-Ambrose, T., 2017. Sex differences in exercise efficacy to improve cognition: A systematic review and meta-analysis of randomized controlled trials in older humans. Front. Neuroendocrinol. 46, 71–85. doi:10.1016/j.yfrne.2017.04.002

Barha, C.K., Hsu, C.-L., Ten Brinke, L., Liu-Ambrose, T., 2019. Biological sex: A potential moderator of physical activity efficacy on brain health. Front. Aging Neurosci. 11, 329. doi:10.3389/fnagi.2019.00329

Barha, C.K., Liu-Ambrose, T., 2018. Exercise and the aging brain: considerations for sex differences. Brain Plast. 4, 53–63. doi:10.3233/BPL-180067

Bassett, D.S., Bullmore, E., 2006. Small-world brain networks. Neuroscientist 12, 512–523. doi:10.1177/1073858406293182

Bastos, A.M., Schoffelen, J.-M., 2015. A Tutorial Review of Functional Connectivity Analysis Methods and Their Interpretational Pitfalls. Front. Syst. Neurosci. 9, 175. doi:10.3389/fnsys.2015.00175

Behzadi, Y., Restom, K., Liau, J., Liu, T.T., 2007. A component based noise correction method (CompCor) for BOLD and perfusion based fMRI. Neuroimage 37, 90–101. doi:10.1016/j.neuroimage.2007.04.042

Belleville, S., Bherer, L., 2012. Biomarkers of cognitive training effects in aging. Curr. Transl. Geriatr. Exp. Gerontol. Rep. 1, 104–110. doi:10.1007/s13670-012-0014-5

Benjamini, Y., Hochberg, Y., 1995. Controlling the false discovery rate: a practical and powerful approach to multiple testing. Journal of the Royal Statistical Society: Series B (Methodological) 57, 289–300. doi:10.1111/j.2517-6161.1995.tb02031.x

Betzel, R.F., Byrge, L., He, Y., Goñi, J., Zuo, X.-N., Sporns, O., 2014. Changes in structural and functional connectivity among resting-state networks across the human lifespan. Neuroimage 102 Pt 2, 345–357. doi:10.1016/j.neuroimage.2014.07.067

Biswal, B., Yetkin, F.Z., Haughton, V.M., Hyde, J.S., 1995. Functional connectivity in the motor cortex of resting human brain using echo-planar MRI. Magn. Reson. Med. 34, 537–541. doi:10.1002/mrm.1910340409

Borg, G.A., 1982. Psychophysical bases of perceived exertion. Med. Sci. Sports Exerc. 14, 377–381. doi:10.1249/00005768-198205000-00012

Buckner, R.L., 2004. Memory and executive function in aging and AD: multiple factors that cause decline and reserve factors that compensate. Neuron 44, 195–208. doi:10.1016/j.neuron.2004.09.006

Bullmore, E., Sporns, O., 2009. Complex brain networks: Graph theoretical analysis of structural and functional systems. Nat. Rev. Neurosci. 10, 186–198. doi:10.1038/nrn2575

Burdette, J.H., Laurienti, P.J., Espeland, M.A., Morgan, A., Telesford, Q., Vechlekar, C.D., Hayasaka, S., Jennings, J.M., Katula, J.A., Kraft, R.A., Rejeski, W.J., 2010. Using network science to evaluate exercise-associated brain changes in older adults. Front. Aging Neurosci. 2, 23. doi:10.3389/fnagi.2010.00023

Cao, W., Cao, X., Hou, C., Li, T., Cheng, Y., Jiang, L., Luo, C., Li, C., Yao, D., 2016. Effects of Cognitive Training on Resting-State Functional Connectivity of Default Mode, Salience, and Central Executive Networks. Front. Aging Neurosci. 8, 70. doi:10.3389/fnagi.2016.00070

Castells-Sánchez, A., Roig-Coll, F., Dacosta-Aguayo, R., Lamonja-Vicente, N., Sawicka, A.K., Torán-Monserrat, P., Pera, G., Montero-Alía, P., Heras-Tebar, A., Domènech, S., Via, M., Erickson, K.I., Mataró, M., 2021. Exercise and Fitness Neuroprotective Effects: Molecular, Brain Volume and Psychological Correlates and Their Mediating Role in Healthy Late-Middle-Aged Women and Men. Front. Aging Neurosci. 13, 615247. doi:10.3389/fnagi.2021.615247

Castells-Sánchez, A., Roig-Coll, F., Dacosta-Aguayo, R., Lamonja-Vicente, N., Torán-Monserrat, P., Pera, G., García-Molina, A., Tormos, J.M., Montero-Alía, P., Heras-Tébar, A., Soriano-Raya, J.J., Cáceres, C., Domènech, S., Via, M., Erickson, K.I., Mataró, M., 2022. Molecular and Brain Volume Changes Following Aerobic Exercise, Cognitive and Combined Training in Physically Inactive Healthy Late-Middle-Aged Adults: The Projecte Moviment Randomized Controlled Trial. Front. Hum. Neurosci. 16, 854175. doi:10.3389/fnhum.2022.854175

Castells-Sánchez, A., Roig-Coll, F., Lamonja-Vicente, N., Altés-Magret, M., Torán-Monserrat, P., Via, M., García-Molina, A., Tormos, J.M., Heras, A., Alzamora, M.T., Forés, R., Pera, G., Dacosta-Aguayo, R., Soriano-Raya, J.J., Cáceres, C., Montero-Alía, P., Montero-Alía, J.J., Jimenez-Gonzalez, M.M., Hernández-Pérez, M., Perera, A., Mataró, M., 2019. Effects and mechanisms of cognitive, aerobic exercise, and combined training on cognition, health, and brain outcomes in physically inactive older adults: the projecte moviment protocol. Front. Aging Neurosci. 11, 216. doi:10.3389/fnagi.2019.00216

Chaddock-Heyman, L., Erickson, K.I., Holtrop, J.L., Voss, M.W., Pontifex, M.B., Raine, L.B., Hillman, C.H., Kramer, A.F., 2014. Aerobic fitness is associated with greater white matter integrity in children. Front. Hum. Neurosci. 8, 584. doi:10.3389/fnhum.2014.00584

Chan, M.Y., Alhazmi, F.H., Park, D.C., Savalia, N.K., Wig, G.S., 2017. Resting-State Network Topology Differentiates Task Signals across the Adult Life Span. J. Neurosci. 37, 2734–2745. doi:10.1523/JNEUROSCI.2406-16.2017

Chapman, S.B., Aslan, S., Spence, J.S., Defina, L.F., Keebler, M.W., Didehbani, N., Lu, H., 2013. Shorter term aerobic exercise improves brain, cognition, and cardiovascular fitness in aging. Front. Aging Neurosci. 5, 75. doi:10.3389/fnagi.2013.00075

Chapman, S.B., Aslan, S., Spence, J.S., Hart, J.J., Bartz, E.K., Didehbani, N., Keebler, M.W., Gardner, C.M., Strain, J.F., DeFina, L.F., Lu, H., 2015. Neural mechanisms of brain plasticity with complex cognitive training in healthy seniors. Cereb. Cortex 25, 396–405. doi:10.1093/cercor/bht234

Chen, F.-T., Hopman, R.J., Huang, C.-J., Chu, C.-H., Hillman, C.H., Hung, T.-M., Chang, Y.-K., 2020. The Effect of Exercise Training on Brain Structure and Function in Older Adults: A Systematic Review Based on Evidence from Randomized Control Trials. J. Clin. Med. 9. doi:10.3390/jcm9040914

Chirles, T.J., Reiter, K., Weiss, L.R., Alfini, A.J., Nielson, K.A., Smith, J.C., 2017. Exercise training and functional connectivity changes in mild cognitive impairment and healthy elders. J Alzheimers Dis 57, 845–856. doi:10.3233/JAD-161151

Colcombe, S.J., Kramer, A.F., 2003. Fitness effects on the cognitive function of older adults: a meta-analytic study. Psychol. Sci. 14, 125–130. doi:10.1111/1467-9280.t01-1-01430

Craik, F.I.M., Bialystok, E., 2006. Cognition through the lifespan: mechanisms of change. Trends Cogn Sci (Regul Ed) 10, 131–138. doi:10.1016/j.tics.2006.01.007

Craik, F.I.M., Byrd, M., 1982. Aging and cognitive deficits, in: Craik, F I M, Trehub, S. (Eds.), Aging and Cognitive Processes. Springer US, Boston, MA, pp. 191–211. doi:10.1007/978-1-4684-4178-9_11

Damoiseaux, J.S., Beckmann, C.F., Arigita, E.J.S., Barkhof, F., Scheltens, P., Stam, C.J., Smith, S.M., Rombouts, S.A.R.B., 2008. Reduced resting-state brain activity in the “default network” in normal aging. Cereb. Cortex 18, 1856–1864. doi:10.1093/cercor/bhm207

Damoiseaux, J.S., Rombouts, S.A.R.B., Barkhof, F., Scheltens, P., Stam, C.J., Smith, S.M., Beckmann, C.F., 2006. Consistent resting-state networks across healthy subjects. Proc Natl Acad Sci USA 103, 13848–13853. doi:10.1073/pnas.0601417103

Damoiseaux, J.S., 2017. Effects of aging on functional and structural brain connectivity. Neuroimage 160, 32–40. doi:10.1016/j.neuroimage.2017.01.077

de Oliveira Rosa, V., Rosa Franco, A., Abrahão Salum Júnior, G., Moreira-Maia, C.R., Wagner, F., Simioni, A., de Fraga Bassotto, C., R Moritz, G., Schaffer Aguzzoli, C., Buchweitz, A., Schmitz, M., Rubia, K., Paim Rohde, L.A., 2020. Effects of computerized cognitive training as add-on treatment to stimulants in ADHD: a pilot fMRI study. Brain Imaging Behav. 14, 1933–1944. doi:10.1007/s11682-019-00137-0

Deng, L., Cheng, Y., Cao, X., Feng, W., Zhu, H., Jiang, L., Wu, W., Tong, S., Sun, J., Li, C., 2019. The effect of cognitive training on the brain’s local connectivity organization in healthy older adults. Sci. Rep. 9, 9033. doi:10.1038/s41598-019-45463-x

Dennis, E.L., Thompson, P.M., 2014. Functional brain connectivity using fMRI in aging and Alzheimer’s disease. Neuropsychol. Rev. 24, 49–62. doi:10.1007/s11065-014-9249-6

Dimitriadis, S.I., Lyssoudis, C., Tsolaki, A.C., Lazarou, E., Kozori, M., Tsolaki, M., 2021. Greek High Phenolic Early Harvest Extra Virgin Olive Oil Reduces the Over-Excitation of Information-Flow Based on Dominant Coupling Mode (DoCM) Model in Patients with Mild Cognitive Impairment: An EEG Resting-State Validation Approach. J Alzheimers Dis 83, 191–207. doi:10.3233/JAD-210454

Dimitriadis, S.I., 2022. Assessing the Repeatability of Multi-Frequency Multi-Layer Brain Network Topologies Across Alternative Researcher’s Choice Paths. Neuroinformatics. doi:10.1007/s12021-022-09610-6

Edde, M., Dilharreguy, B., Theaud, G., Chanraud, S., Helmer, C., Dartigues, J.-F., Amieva, H., Allard, M., Descoteaux, M., Catheline, G., 2020. Age-related change in episodic memory: role of functional and structural connectivity between the ventral posterior cingulate and the parietal cortex. Brain Struct. Funct. 225, 2203–2218. doi:10.1007/s00429-020-02121-7

Edde, M., Leroux, G., Altena, E., Chanraud, S., 2021. Functional brain connectivity changes across the human life span: From fetal development to old age. J. Neurosci. Res. 99, 236–262. doi:10.1002/jnr.24669

Eggenberger, P., Schumacher, V., Angst, M., Theill, N., de Bruin, E.D., 2015. Does multicomponent physical exercise with simultaneous cognitive training boost cognitive performance in older adults? A 6-month randomized controlled trial with a 1-year follow-up. Clin. Interv. Aging 10, 1335–1349. doi:10.2147/CIA.S87732

Erickson, K.I., Leckie, R.L., Weinstein, A.M., 2014. Physical activity, fitness, and gray matter volume. Neurobiol. Aging 35, 20–28. doi:10.1016/j.neurobiolaging.2014.03.034

Erickson, K.I., Miller, D.L., Roecklein, K.A., 2012. The aging hippocampus: interactions between exercise, depression, and BDNF. Neuroscientist 18, 82–97. doi:10.1177/1073858410397054

Etnier, J.L., Chang, Y.-K., 2009. The effect of physical activity on executive function: a brief commentary on definitions, measurement issues, and the current state of the literature. J. Sport Exerc. Psychol. 31, 469–483. doi:10.1123/jsep.31.4.469

Fabel, K., Wolf, S.A., Ehninger, D., Babu, H., Leal-Galicia, P., Kempermann, G., 2009. Additive effects of physical exercise and environmental enrichment on adult hippocampal neurogenesis in mice. Front. Neurosci. 3, 50. doi:10.3389/neuro.22.002.2009

Fabre, C., Chamari, K., Mucci, P., Massé-Biron, J., Préfaut, C., 2002. Improvement of cognitive function by mental and/or individualized aerobic training in healthy elderly subjects. Int. J. Sports Med. 23, 415–421. doi:10.1055/s-2002-33735

Fan, Y.-T., Fang, Y.-W., Chen, Y.-P., Leshikar, E.D., Lin, C.-P., Tzeng, O.J.L., Huang, H.-W., Huang, C.-M., 2019. Aging, cognition, and the brain: effects of age-related variation in white matter integrity on neuropsychological function. Aging Ment. Health 23, 831–839. doi:10.1080/13607863.2018.1455804

Farràs-Permanyer, L., Guàrdia-Olmos, J., Peró-Cebollero, M., 2015. Mild cognitive impairment and fMRI studies of brain functional connectivity: the state of the art. Front. Psychol. 6, 1095. doi:10.3389/fpsyg.2015.01095

Flodin, P., Jonasson, L.S., Riklund, K., Nyberg, L., Boraxbekk, C.J., 2017. Does Aerobic Exercise Influence Intrinsic Brain Activity? An Aerobic Exercise Intervention among Healthy Old Adults. Front. Aging Neurosci. 9, 267. doi:10.3389/fnagi.2017.00267

Fox, M.D., Raichle, M.E., 2007. Spontaneous fluctuations in brain activity observed with functional magnetic resonance imaging. Nat. Rev. Neurosci. 8, 700–711. doi:10.1038/nrn2201

Geerligs, L., Renken, R.J., Saliasi, E., Maurits, N.M., Lorist, M.M., 2015. A Brain-Wide Study of Age-Related Changes in Functional Connectivity. Cereb. Cortex 25, 1987–1999. doi:10.1093/cercor/bhu012

Glisky, E.L., 2007. Changes in Cognitive Function in Human Aging, in: Riddle, D.R. (Ed.), Brain Aging: Models, Methods, and Mechanisms, Chapter 1. CRC Press/Taylor & Francis, Boca Raton (FL). doi:10.1201/9781420005523.sec1

Goh, J.O., An, Y., Resnick, S.M., 2012. Differential trajectories of age-related changes in components of executive and memory processes. Psychol. Aging 27, 707–719. doi:10.1037/a0026715

Golden, C.J., 2001. Stroop.Test de Colores y Palabras, 3rd ed. TEA Ediciones., MADRID.

Goodglass, H., Kaplan, E., Barresi, B., 2001. Testde Boston Para el Diagnóstico de la Afasia, 3rd ed. Editorial Médica Panamericana., MADRID.

Grady, C., Sarraf, S., Saverino, C., Campbell, K., 2016. Age differences in the functional interactions among the default, frontoparietal control, and dorsal attention networks. Neurobiol. Aging 41, 159–172. doi:10.1016/j.neurobiolaging.2016.02.020

Grady, C.L., McIntosh, A.R., Beig, S., Keightley, M.L., Burian, H., Black, S.E., 2003. Evidence from functional neuroimaging of a compensatory prefrontal network in Alzheimer’s disease. J. Neurosci. 23, 986–993. doi:10.1523/JNEUROSCI.23-03-00986.2003

Hafkemeijer, A., van der Grond, J., Rombouts, S.A.R.B., 2012. Imaging the default mode network in aging and dementia. Biochim. Biophys. Acta 1822, 431–441. doi:10.1016/j.bbadis.2011.07.008

Harada, C.N., Natelson Love, M.C., Triebel, K.L., 2013. Normal cognitive aging. Clin. Geriatr. Med. 29, 737–752. doi:10.1016/j.cger.2013.07.002

Hasher, L., Zacks, R.T., 1988. Working Memory, Comprehension, and Aging: A Review and a New View, in: Psychology of Learning and Motivation. Elsevier, pp. 193–225. doi:10.1016/S0079-7421(08)60041-9

Hausman, H.K., O’Shea, A., Kraft, J.N., Boutzoukas, E.M., Evangelista, N.D., Van Etten, E.J., Bharadwaj, P.K., Smith, S.G., Porges, E., Hishaw, G.A., Wu, S., DeKosky, S., Alexander, G.E., Marsiske, M., Cohen, R., Woods, A.J., 2020. The Role of Resting-State Network Functional Connectivity in Cognitive Aging. Front. Aging Neurosci. 12, 177. doi:10.3389/fnagi.2020.00177

Hayes, A.F., Rockwood, N.J., 2016. Regression-based statistical mediation and moderation analysis in clinical research: Observations, recommendations, and implementation. Behav. Res. Ther. 98, 39–57. doi:10.1016/j.brat.2016.11.001

He, L., Wang, X., Zhuang, K., Qiu, J., 2020. Decreased Dynamic Segregation but Increased Dynamic Integration of the Resting-state Functional Networks During Normal Aging. Neuroscience 437, 54–63. doi:10.1016/j.neuroscience.2020.04.030

Huang, C.-C., Hsieh, W.-J., Lee, P.-L., Peng, L.-N., Liu, L.-K., Lee, W.-J., Huang, J.-K., Chen, L.-K., Lin, C.-P., 2015. Age-related changes in resting-state networks of a large sample size of healthy elderly. CNS Neurosci. Ther. 21, 817–825. doi:10.1111/cns.12396

Huntley, J.D., Hampshire, A., Bor, D., Owen, A., Howard, R.J., 2017. Adaptive working memory strategy training in early Alzheimer’s disease: randomised controlled trial. Br. J. Psychiatry 210, 61–66. doi:10.1192/bjp.bp.116.182048

Jones, D.T., Machulda, M.M., Vemuri, P., McDade, E.M., Zeng, G., Senjem, M.L., Gunter, J.L., Przybelski, S.A., Avula, R.T., Knopman, D.S., Boeve, B.F., Petersen, R.C., Jack, C.R., 2011. Age-related changes in the default mode network are more advanced in Alzheimer disease. Neurology 77, 1524–1531. doi:10.1212/WNL.0b013e318233b33d

Joubert, C., Chainay, H., 2018. Aging brain: the effect of combined cognitive and physical training on cognition as compared to cognitive and physical training alone - a systematic review. Clin. Interv. Aging 13, 1267–1301. doi:10.2147/CIA.S165399

Kaiser, A., Reneman, L., Solleveld, M.M., Coolen, B.F., Scherder, E.J.A., Knutsson, L., Bjørnerud, A., van Osch, M.J.P., Wijnen, J.P., Lucassen, P.J., Schrantee, A., 2021. A Randomized Controlled Trial on the Effects of a 12-Week High- vs. Low-Intensity Exercise Intervention on Hippocampal Structure and Function in Healthy, Young Adults. Front. Psychiatry 12, 780095. doi:10.3389/fpsyt.2021.780095

Kang, W., Wang, J., Malvaso, A., 2021. Inhibitory Control in Aging: The Compensation-Related Utilization of Neural Circuits Hypothesis. Front. Aging Neurosci. 13, 771885. doi:10.3389/fnagi.2021.771885

Kim, H., Chey, J., Lee, S., 2017. Effects of multicomponent training of cognitive control on cognitive function and brain activation in older adults. Neurosci. Res. 124, 8–15. doi:10.1016/j.neures.2017.05.004

King, B.R., van Ruitenbeek, P., Leunissen, I., Cuypers, K., Heise, K.F., Santos Monteiro, T., Hermans, L., Levin, O., Albouy, G., Mantini, D., Swinnen, S.P., 2018. Age-Related Declines in Motor Performance are Associated With Decreased Segregation of Large-Scale Resting State Brain Networks. Cereb. Cortex 28, 4390–4402. doi:10.1093/cercor/bhx297

Kline, G.M., Porcari, J.P., Hintermeister, R., Freedson, P.S., Ward, A., McCarron, R.F., Ross, J., Rippe, J.M., 1987. Estimation of VO2max from a one-mile track walk, gender, age, and body weight. Med. Sci. Sports Exerc. 19, 253–259.

Kraft, E., 2012. Cognitive function, physical activity, and aging: possible biological links and implications for multimodal interventions. Neuropsychol. Dev. Cogn. B Aging Neuropsychol. Cogn. 19, 248–263. doi:10.1080/13825585.2011.645010

Kramer, A.F., Humphrey, D.G., Larish, J.F., Logan, G.D., Strayer, D.L., 1994. Aging and inhibition: beyond a unitary view of inhibitory processing in attention. Psychol. Aging 9, 491–512. doi:10.1037/0882-7974.9.4.491

Lampit, A., Hallock, H., Valenzuela, M., 2014. Computerized cognitive training in cognitively healthy older adults: a systematic review and meta-analysis of effect modifiers. PLoS Med. 11, e1001756. doi:10.1371/journal.pmed.1001756

Lampit, A., Valenzuela, M., Gates, N.J., 2015. Computerized cognitive training is beneficial for older adults. J. Am. Geriatr. Soc. 63, 2610–2612. doi:10.1111/jgs.13825

Lezak, M.D., Howieson, D.B., Bigler, E.D., Tranel, D., 2012. Neuropsychological Assessment, 5th ed. Oxford University Press, Oxford.

Li, R., Zhu, X., Yin, S., Niu, Y., Zheng, Z., Huang, X., Wang, B., Li, J., 2014. Multimodal intervention in older adults improves resting-state functional connectivity between the medial prefrontal cortex and medial temporal lobe. Front. Aging Neurosci. 6, 39. doi:10.3389/fnagi.2014.00039

Li, T., Yao, Y., Cheng, Y., Xu, B., Cao, X., Waxman, D., Feng, W., Shen, Y., Li, Q., Wang, J., Wu, W., Li, C., Feng, J., 2016. Cognitive training can reduce the rate of cognitive aging: a neuroimaging cohort study. BMC Geriatr. 16, 12. doi:10.1186/s12877-016-0194-5

Lövdén, M., Brehmer, Y., Li, S.-C., Lindenberger, U., 2012. Training-induced compensation versus magnification of individual differences in memory performance. Front. Hum. Neurosci. 6, 141. doi:10.3389/fnhum.2012.00141

Luo, C., Zhang, X., Cao, X., Gan, Y., Li, T., Cheng, Y., Cao, W., Jiang, L., Yao, D., Li, C., 2016. The lateralization of intrinsic networks in the aging brain implicates the effects of cognitive training. Front. Aging Neurosci. 8, 32. doi:10.3389/fnagi.2016.00032

Luppi, A.I., Gellersen, H.M., Peattie, A.R.D., Manktelow, A.E., Menon, D., Dimitriadis, S.I., Stamatakis, E.A., 2021. Searching for consistent brain network topologies across the garden of (shortest) forking paths. BioRxiv. doi:10.1101/2021.07.13.452257

Maass, A., Düzel, S., Goerke, M., Becke, A., Sobieray, U., Neumann, K., Lövden, M., Lindenberger, U., Bäckman, L., Braun-Dullaeus, R., Ahrens, D., Heinze, H.J., Müller, N.G., Düzel, E., 2015. Vascular hippocampal plasticity after aerobic exercise in older adults. Mol. Psychiatry 20, 585–593. doi:10.1038/mp.2014.114

Madden, D.J., Jain, S., Monge, Z.A., Cook, A.D., Lee, A., Huang, H., Howard, C.M., Cohen, J.R., 2020. Influence of structural and functional brain connectivity on age-related differences in fluid cognition. Neurobiol. Aging 96, 205–222. doi:10.1016/j.neurobiolaging.2020.09.010

Mancho-Fora, N., Montalà-Flaquer, M., Farràs-Permanyer, L., Zarabozo-Hurtado, D., Gallardo-Moreno, G.B., Gudayol-Farré, E., Peró-Cebollero, M., Guàrdia-Olmos, J., 2020. Network change point detection in resting-state functional connectivity dynamics of mild cognitive impairment patients. Int. J. Clin. Health Psychol. 20, 200–212. doi:10.1016/j.ijchp.2020.07.005

McFadden, K.L., Cornier, M.-A., Melanson, E.L., Bechtell, J.L., Tregellas, J.R., 2013. Effects of exercise on resting-state default mode and salience network activity in overweight/obese adults. Neuroreport 24, 866–871. doi:10.1097/WNR.0000000000000013

McGregor, K.M., Crosson, B., Krishnamurthy, L.C., Krishnamurthy, V., Hortman, K., Gopinath, K., Mammino, K.M., Omar, J., Nocera, J.R., 2018. Effects of a 12-Week Aerobic Spin Intervention on Resting State Networks in Previously Sedentary Older Adults. Front. Psychol. 9, 2376. doi:10.3389/fpsyg.2018.02376

Mevel, K., Chételat, G., Eustache, F., Desgranges, B., 2011. The default mode network in healthy aging and Alzheimer’s disease. Int J Alzheimers Dis 2011, 535816. doi:10.4061/2011/535816

Northey, J.M., Cherbuin, N., Pumpa, K.L., Smee, D.J., Rattray, B., 2018. Exercise interventions for cognitive function in adults older than 50: a systematic review with meta-analysis. Br. J. Sports Med. 52, 154–160. doi:10.1136/bjsports-2016-096587

Olson, A.K., Eadie, B.D., Ernst, C., Christie, B.R., 2006. Environmental enrichment and voluntary exercise massively increase neurogenesis in the adult hippocampus via dissociable pathways. Hippocampus 16, 250–260. doi:10.1002/hipo.20157

Onoda, K., Ishihara, M., Yamaguchi, S., 2012. Decreased functional connectivity by aging is associated with cognitive decline. J. Cogn. Neurosci. 24, 2186–2198. doi:10.1162/jocn_a_00269

Park, D.C., Lautenschlager, G., Hedden, T., Davidson, N.S., Smith, A.D., Smith, P.K., 2002. Models of visuospatial and verbal memory across the adult life span. Psychol. Aging 17, 299–320. doi:10.1037//0882-7974.17.2.299

Peña-Casanova, J., Quiñones-Ubeda, S., Gramunt-Fombuena, N., Quintana-Aparicio, M., Aguilar, M., Badenes, D., Cerulla, N., Molinuevo, J.L., Ruiz, E., Robles, A., Barquero, M.S., Antúnez, C., Martínez-Parra, C., Frank-García, A., Fernández, M., Alfonso, V., Sol, J.M., Blesa, R., NEURONORMA Study Team, 2009. Spanish Multicenter Normative Studies (NEURONORMA Project): norms for verbal fluency tests. Arch. Clin. Neuropsychol. 24, 395–411. doi:10.1093/arclin/acp042

Pieramico, V., Esposito, R., Sensi, F., Cilli, F., Mantini, D., Mattei, P.A., Frazzini, V., Ciavardelli, D., Gatta, V., Ferretti, A., Romani, G.L., Sensi, S.L., 2012. Combination training in aging individuals modifies functional connectivity and cognition, and is potentially affected by dopamine-related genes. PLoS ONE 7, e43901. doi:10.1371/journal.pone.0043901

Prehn, K., Lesemann, A., Krey, G., Witte, A.V., Köbe, T., Grittner, U., Flöel, A., 2019. Using resting-state fMRI to assess the effect of aerobic exercise on functional connectivity of the DLPFC in older overweight adults. Brain Cogn. 131, 34–44. doi:10.1016/j.bandc.2017.08.006

Raz, N., 2001. Ageing and the brain, in: John Wiley & Sons, Ltd (Ed.), ELS. Wiley, pp. 1–8. doi:10.1002/9780470015902.a0003375.pub3

Reuter-Lorenz, P.A., Lustig, C., 2005. Brain aging: reorganizing discoveries about the aging mind. Curr. Opin. Neurobiol. 15, 245–251. doi:10.1016/j.conb.2005.03.016

Rey, A., 2009. REY. Test deCopia de una Figura Compleja, 3rd ed. TEA Ediciones., MADRID.

Ritchie, S.J., Dickie, D.A., Cox, S.R., Valdes Hernandez, M.D.C., Corley, J., Royle, N.A., Pattie, A., Aribisala, B.S., Redmond, P., Muñoz Maniega, S., Taylor, A.M., Sibbett, R., Gow, A.J., Starr, J.M., Bastin, M.E., Wardlaw, J.M., Deary, I.J., 2015. Brain volumetric changes and cognitive ageing during the eighth decade of life. Hum. Brain Mapp. 36, 4910–4925. doi:10.1002/hbm.22959

Roig-Coll, F., Castells-Sánchez, A., Lamonja-Vicente, N., Torán-Monserrat, P., Pera, G., García-Molina, A., Tormos, J.M., Montero-Alía, P., Alzamora, M.T., Dacosta-Aguayo, R., Soriano-Raya, J.J., Cáceres, C., Erickson, K.I., Mataró, M., 2020. Effects of Aerobic Exercise, Cognitive and Combined Training on Cognition in Physically Inactive Healthy Late-Middle-Aged Adults: The Projecte Moviment Randomized Controlled Trial. Front. Aging Neurosci. 12, 590168. doi:10.3389/fnagi.2020.590168

Ross, L.A., Webb, C.E., Whitaker, C., Hicks, J.M., Schmidt, E.L., Samimy, S., Dennis, N.A., Visscher, K.M., 2019. The effects of useful field of view training on brain activity and connectivity. J. Gerontol. B Psychol. Sci. Soc. Sci. 74, 1152–1162. doi:10.1093/geronb/gby041

Ruiz Comellas, A., Pera, G., Baena Díez, J.M., Mundet Tudurí, X., Alzamora Sas, T., Elosua, R., Torán Monserrat, P., Heras, A., Forés Raurell, R., Fusté Gamisans, M., Fàbrega Camprubí, M., 2012. [validation of a spanish short version of the minnesota leisure time physical activity questionnaire (VREM)]. Rev. Esp. Salud Publica 86, 495–508. doi:10.4321/S1135-57272012000500004

Sala-Llonch, R., Bartrés-Faz, D., Junqué, C., 2015. Reorganization of brain networks in aging: a review of functional connectivity studies. Front. Psychol. 6, 663. doi:10.3389/fpsyg.2015.00663

Salthouse, T., Maurer, T., 1996. Aging, job performance and career development, in: Handbook of the Psychology of Aging. Academic, San Francisco, CA, pp. 353–364.

Salthouse, T.A., 1994. Age-related differences in basic cognitive processes: implications for work. Exp. Aging Res. 20, 249–255. doi:10.1080/03610739408253974

Salthouse, T.A., 2010. Major Issues in Cognitive Aging, 1st ed. Oxford Univ. Press, New York.

Schmidt, M., 1996. Rey Auditory VerbalLearning Test: A Handbook, 1st ed. CA: Western Psychological Services., Los Angeles.

Shao, Y., Mang, J., Li, P., Wang, J., Deng, T., Xu, Z., 2015. Computer-Based Cognitive Programs for Improvement of Memory, Processing Speed and Executive Function during Age-Related Cognitive Decline: A Meta-Analysis. PLoS ONE 10, e0130831. doi:10.1371/journal.pone.0130831

Shatil, E., 2013. Does combined cognitive training and physical activity training enhance cognitive abilities more than either alone? A four-condition randomized controlled trial among healthy older adults. Front. Aging Neurosci. 5, 8. doi:10.3389/fnagi.2013.00008

Shaw, E.E., Schultz, A.P., Sperling, R.A., Hedden, T., 2015. Functional Connectivity in Multiple Cortical Networks Is Associated with Performance Across Cognitive Domains in Older Adults. Brain Connect. 5, 505–516. doi:10.1089/brain.2014.0327

Shehzad, Z., Kelly, A.M.C., Reiss, P.T., Gee, D.G., Gotimer, K., Uddin, L.Q., Lee, S.H., Margulies, D.S., Roy, A.K., Biswal, B.B., Petkova, E., Castellanos, F.X., Milham, M.P., 2009. The resting brain: unconstrained yet reliable. Cereb. Cortex 19, 2209–2229. doi:10.1093/cercor/bhn256

Siman-Tov, T., Bosak, N., Sprecher, E., Paz, R., Eran, A., Aharon-Peretz, J., Kahn, I., 2016. Early Age-Related Functional Connectivity Decline in High-Order Cognitive Networks. Front. Aging Neurosci. 8, 330. doi:10.3389/fnagi.2016.00330

Song, J., Birn, R.M., Boly, M., Meier, T.B., Nair, V.A., Meyerand, M.E., Prabhakaran, V., 2014. Age-related reorganizational changes in modularity and functional connectivity of human brain networks. Brain Connect. 4, 662–676. doi:10.1089/brain.2014.0286

Spreen, O., 1998. A compendium of neuropsychological tests: Administration, norms, and commentary, 2nd ed. Oxford University Press., OXFORD.

Stillman, C.M., Cohen, J., Lehman, M.E., Erickson, K.I., 2016. Mediators of physical activity on neurocognitive function: A review at multiple levels of analysis. Front. Hum. Neurosci. 10, 626. doi:10.3389/fnhum.2016.00626

Stillman, C.M., Donofry, S.D., Erickson, K.I., 2019. Exercise, fitness and the aging brain: A review of functional connectivity in aging. AOP 3. doi:10.31296/aop.v3i4.98

Stillman, C.M., Esteban-Cornejo, I., Brown, B., Bender, C.M., Erickson, K.I., 2020. Effects of exercise on brain and cognition across age groups and health states. Trends Neurosci. 43, 533–543. doi:10.1016/j.tins.2020.04.010

Stillman, C.M., Uyar, F., Huang, H., Grove, G.A., Watt, J.C., Wollam, M.E., Erickson, K.I., 2018. Cardiorespiratory fitness is associated with enhanced hippocampal functional connectivity in healthy young adults. Hippocampus 28, 239–247. doi:10.1002/hipo.22827

Strenziok, M., Parasuraman, R., Clarke, E., Cisler, D.S., Thompson, J.C., Greenwood, P.M., 2014. Neurocognitive enhancement in older adults: comparison of three cognitive training tasks to test a hypothesis of training transfer in brain connectivity. Neuroimage 85 Pt 3, 1027–1039. doi:10.1016/j.neuroimage.2013.07.069

Székely, G.J., Rizzo, M.L., 2013. The distance correlation-test of independence in high dimension. J. Multivar. Anal. 117, 193–213. doi:10.1016/j.jmva.2013.02.012

Takeuchi, H., Magistro, D., Kotozaki, Y., Motoki, K., Nejad, K.K., Nouchi, R., Jeong, H., Sato, C., Sessa, S., Nagatomi, R., Zecca, M., Takanishi, A., Kawashima, R., 2020. Effects of Simultaneously Performed Dual-Task Training with Aerobic Exercise and Working Memory Training on Cognitive Functions and Neural Systems in the Elderly. Neural Plast. 2020, 1–17. doi:10.1155/2020/3859824

Tao, J., Liu, J., Chen, X., Xia, R., Li, M., Huang, M., Li, S., Park, J., Wilson, G., Lang, C., Xie, G., Zhang, B., Zheng, G., Chen, L., Kong, J., 2019. Mind-body exercise improves cognitive function and modulates the function and structure of the hippocampus and anterior cingulate cortex in patients with mild cognitive impairment. Neuroimage Clin. 23, 101834. doi:10.1016/j.nicl.2019.101834

Tao, J., Liu, J., Egorova, N., Chen, X., Sun, S., Xue, X., Huang, J., Zheng, G., Wang, Q., Chen, L., Kong, J., 2016. Increased Hippocampus-Medial Prefrontal Cortex Resting-State Functional Connectivity and Memory Function after Tai Chi Chuan Practice in Elder Adults. Front. Aging Neurosci. 8, 25. doi:10.3389/fnagi.2016.00025

Taya, F., Sun, Y., Babiloni, F., Thakor, N., Bezerianos, A., 2015. Brain enhancement through cognitive training: a new insight from brain connectome. Front. Syst. Neurosci. 9, 44. doi:10.3389/fnsys.2015.00044

Ten Brinke, L., Lee, J.J., Carney, D.R., 2019. Different physiological reactions when observing lies versus truths: Initial evidence and an intervention to enhance accuracy. J. Pers. Soc. Psychol. 117, 560–578. doi:10.1037/pspi0000175

Ten Brinke, L.F., Davis, J.C., Barha, C.K., Liu-Ambrose, T., 2017. Effects of computerized cognitive training on neuroimaging outcomes in older adults: a systematic review. BMC Geriatr. 17, 139. doi:10.1186/s12877-017-0529-x

Tomasi, D., Volkow, N.D., 2012. Aging and functional brain networks. Mol. Psychiatry 17, 471, 549–58. doi:10.1038/mp.2011.81

Tombaugh, T., 2004. Trail Making Test A and B: Normative data stratified by age and education. Arch. Clin. Neuropsychol. 19, 203–214. doi:10.1016/S0887-6177(03)00039-8

Tozzi, L., Carballedo, A., Lavelle, G., Doolin, K., Doyle, M., Amico, F., McCarthy, H., Gormley, J., Lord, A., O’Keane, V., Frodl, T., 2016. Longitudinal functional connectivity changes correlate with mood improvement after regular exercise in a dose-dependent fashion. Eur. J. Neurosci. 43, 1089–1096. doi:10.1111/ejn.13222

Tzourio-Mazoyer, N., Landeau, B., Papathanassiou, D., Crivello, F., Etard, O., Delcroix, N., Mazoyer, B., Joliot, M., 2002. Automated anatomical labeling of activations in SPM using a macroscopic anatomical parcellation of the MNI MRI single-subject brain. Neuroimage 15, 273–289. doi:10.1006/nimg.2001.0978

van Balkom, T.D., van den Heuvel, O.A., Berendse, H.W., van der Werf, Y.D., Vriend, C., 2020. The effects of cognitive training on brain network activity and connectivity in aging and neurodegenerative diseases: a systematic review. Neuropsychol. Rev. 30, 267–286. doi:10.1007/s11065-020-09440-w

Varangis, E., Habeck, C.G., Razlighi, Q.R., Stern, Y., 2019. The effect of aging on resting state connectivity of predefined networks in the brain. Front. Aging Neurosci. 11, 234. doi:10.3389/fnagi.2019.00234

Veldsman, M., Churilov, L., Werden, E., Li, Q., Cumming, T., Brodtmann, A., 2017. Physical activity after stroke is associated with increased interhemispheric connectivity of the dorsal attention network. Neurorehabil. Neural Repair 31, 157–167. doi:10.1177/1545968316666958

Voss, M.W., Erickson, K.I., Prakash, R.S., Chaddock, L., Malkowski, E., Alves, H., Kim, J.S., Morris, K.S., White, S.M., Wójcicki, T.R., Hu, L., Szabo, A., Klamm, E., McAuley, E., Kramer, A.F., 2010a. Functional connectivity: a source of variance in the association between cardiorespiratory fitness and cognition? Neuropsychologia 48, 1394–1406. doi:10.1016/j.neuropsychologia.2010.01.005

Voss, M.W., Prakash, R.S., Erickson, K.I., Basak, C., Chaddock, L., Kim, J.S., Alves, H., Heo, S., Szabo, A.N., White, S.M., Wójcicki, T.R., Mailey, E.L., Gothe, N., Olson, E.A., McAuley, E., Kramer, A.F., 2010b. Plasticity of brain networks in a randomized intervention trial of exercise training in older adults. Front. Aging Neurosci. 2. doi:10.3389/fnagi.2010.00032

Voss, M.W., Weng, T.B., Burzynska, A.Z., Wong, C.N., Cooke, G.E., Clark, R., Fanning, J., Awick, E., Gothe, N.P., Olson, E.A., McAuley, E., Kramer, A.F., 2016. Fitness, but not physical activity, is related to functional integrity of brain networks associated with aging. Neuroimage 131, 113–125. doi:10.1016/j.neuroimage.2015.10.044

Wang, L., Laviolette, P., O’Keefe, K., Putcha, D., Bakkour, A., Van Dijk, K.R.A., Pihlajamäki, M., Dickerson, B.C., Sperling, R.A., 2010. Intrinsic connectivity between the hippocampus and posteromedial cortex predicts memory performance in cognitively intact older individuals. Neuroimage 51, 910–917. doi:10.1016/j.neuroimage.2010.02.046

Wang, L., Zang, Y., He, Y., Liang, M., Zhang, X., Tian, L., Wu, T., Jiang, T., Li, K., 2006. Changes in hippocampal connectivity in the early stages of Alzheimer’s disease: evidence from resting state fMRI. Neuroimage 31, 496–504. doi:10.1016/j.neuroimage.2005.12.033

Wechsler, D., 2001. WAIS-III. Escalade Inteligencia de Wechsler Para Adultos, 3rd ed, WAIS-III. Escalade Inteligencia de Wechsler Para Adultos. TEA Ediciones, MADRID.

Wecker, N.S., Kramer, J.H., Hallam, B.J., Delis, D.C., 2005. Mental flexibility: age effects on switching. Neuropsychology 19, 345–352. doi:10.1037/0894-4105.19.3.345

Weng, T.B., Pierce, G.L., Darling, W.G., Falk, D., Magnotta, V.A., Voss, M.W., 2017. The Acute Effects of Aerobic Exercise on the Functional Connectivity of Human Brain Networks. Brain Plast. 2, 171–190. doi:10.3233/BPL-160039

Whitfield-Gabrieli, S., Nieto-Castanon, A., 2012. Conn: a functional connectivity toolbox for correlated and anticorrelated brain networks. Brain Connect. 2, 125–141. doi:10.1089/brain.2012.0073

Won, J., Callow, D.D., Pena, G.S., Gogniat, M.A., Kommula, Y., Arnold-Nedimala, N.A., Jordan, L.S., Smith, J.C., 2021a. Evidence for exercise-related plasticity in functional and structural neural network connectivity. Neurosci. Biobehav. Rev. 131, 923–940. doi:10.1016/j.neubiorev.2021.10.013

Won, J., Faroqi-Shah, Y., Callow, D.D., Williams, A., Awoyemi, A., Nielson, K.A., Smith, J.C., 2021b. Association between greater cerebellar network connectivity and improved phonemic fluency performance after exercise training in older adults. Cerebellum 20, 542–555. doi:10.1007/s12311-020-01218-3

Yang, A.C., Tsai, S.-J., Liu, M.-E., Huang, C.-C., Lin, C.-P., 2016. The Association of Aging with White Matter Integrity and Functional Connectivity Hubs. Front. Aging Neurosci. 8, 143. doi:10.3389/fnagi.2016.00143

Yeo, B.T.T., Krienen, F.M., Sepulcre, J., Sabuncu, M.R., Lashkari, D., Hollinshead, M., Roffman, J.L., Smoller, J.W., Zöllei, L., Polimeni, J.R., Fischl, B., Liu, H., Buckner, R.L., 2011. The organization of the human cerebral cortex estimated by intrinsic functional connectivity. J. Neurophysiol. 106, 1125–1165. doi:10.1152/jn.00338.2011

