## Supplementary Material for "Intrinsic functional brain connectivity changes following aerobic exercise, computerized cognitive training and their combination in healthy late-middle-aged adults: the Projecte Moviment"

**Address: Department of Clinical Psychology and psychobiology, University of Barcelona, Passeig Vall d’Hebron 171, 08035 Barcelona, Spain.**

| **Table 1**  Cognitive outcomes: variables and measures | | | |
| --- | --- | --- | --- |
| Composites 1^st^ Level | Composites 2^nd^ Level | Tests - Subtest | Measure |
| Executive Function | Flexibility | TMT B -A | Z score |
|  | Fluency | Letter fluency | Z score |
|  |  | Category fluency | Z score |
|  | Inhibition | Stroop - Interference | Z score |
|  | Working Memory | WAIS III - Backward Span | Z score |
| Visuospatial Function | Visuospatial Function | ROCF - Copy Accuracy | Z score |
| Language | Language | BNT (15 items) | Z score |
| Attention - Speed | Attention | WAIS III - Forward Span | Z score |
|  |  | WAIS III - Digit Symbol Coding | Z score |
|  |  | WAIS-III - Symbol Search | Z score |
|  | Speed | TMT - A | Z score |
|  |  | ROCF - Copy Time | Z score |
| Memory | Visual Memory | ROCF - Memory Accuracy | Z score |
|  | Verbal Memory | RAVLT - Total Learning | Z score |
|  |  | RAVLT - Recall II | Z score |
| Global Cognitive Function | *Sum of all domains* | | Z score |
| BNT, Boston Naming Test (Goodglass et al., 2001); RAVLT, Rey Auditory Verbal Learning Test (Schmidt, 1996); ROCF, Rey-Osterrieth Complex Figure (Rey, 2009); Stroop Test (Golden, 2001); TMT, Trail Making Test (Tombaugh, 2004); Verbal Fluency Tests (Peña-Casanova et al., 2009); WAIS-III, Wechsler Adult Intelligence Scale (Wechsler, 2001). | | | |

| **Table 2.1**  Group comparison at Baseline: z-scores of Cognitive Domains | | | | | |
| --- | --- | --- | --- | --- | --- |
| Variables | Groups | n | Mean | SD | ANOVA / H de Kruskall Wallis |
| Executive Function | AE | 24 | -0.01 | 0.72 | F(3,75) = 0.81, *p* = .492 |
|  | CCT | 23 | 0.16 | 0.60 |  |
|  | COMB | 19 | -0.08 | 0.50 |  |
|  | Control | 13 | -0.15 | 0.81 |  |
| Flexibility | AE | 25 | 0.10 | 1.05 | H(3) = 1.53, *p* = .676 |
|  | CCT | 23 | -0.10 | 1.06 |  |
|  | COMB | 19 | -0.03 | 0.80 |  |
|  | Control | 14 | 0.04 | 1.12 |  |
| Fluency | AE | 25 | 0.02 | 0.91 | F(3,77) = 1.19, *p* = .318 |
|  | CCT | 23 | 0.22 | 0.76 |  |
|  | COMB | 19 | -0.10 | 0.82 |  |
|  | Control | 14 | -0.28 | 0.80 |  |
| Inhibition | AE | 24 | -0.06 | 1.03 | F(3,77) = 1.18, *p* = .323 |
|  | CCT | 23 | 0.32 | 0.86 |  |
|  | COMB | 19 | -0.16 | 1.18 |  |
|  | Control | 15 | -0.20 | 0.89 |  |
| Working Memory | AE | 25 | -0.06 | 1.10 | H(3) = 1.29, *p* = .732 |
|  | CCT | 23 | 0.14 | 1.05 |  |
|  | COMB | 19 | -0.01 | 0.69 |  |
|  | Control | 15 | -0.11 | 1.16 |  |
| Visuospatial Function | AE | 25 | 0.14 | 0.93 | H(3) = 3.40, *p* = .334 |
|  | CCT | 23 | -0.20 | 1.04 |  |
|  | COMB | 19 | -0.19 | 1.24 |  |
|  | Control | 15 | 0.32 | 0.62 |  |
| Language | AE | 25 | 0.01 | 1.09 | H(3) = 0.71, *p* = .870 |
|  | CCT | 23 | -0.04 | 1.08 |  |
|  | COMB | 19 | -0.10 | 0.91 |  |
|  | Control | 15 | 0.16 | 0.89 |  |
| Attention-Speed | AE | 24 | -0.02 | 1.01 | H(3) = 2.34, *p* = .506 |
|  | CCT | 23 | 0.13 | 0.65 |  |
|  | COMB | 19 | -0.17 | 0.68 |  |
|  | Control | 14 | 0.09 | 0.44 |  |
| Attention | AE | 25 | 0.03 | 0.95 | F(3,77) = 0.70, *p* = .553 |
|  | CCT | 23 | 0.15 | 0.71 |  |
|  | COMB | 19 | -0.20 | 0.76 |  |
|  | Control | 14 | 0.03 | 0.62 |  |
| Speed | AE | 24 | -0.12 | 1.21 | H(3) = 1.64, *p* = .650 |
|  | CCT | 23 | 0.11 | 0.77 |  |
|  | COMB | 19 | -0.12 | 0.74 |  |
|  | Control | 15 | 0.17 | 0.32 |  |
| Memory | AE | 25 | 0.03 | 0.70 | H(3) = 0.95, *p* = .815 |
|  | CCT | 23 | 0.13 | 0.66 |  |
|  | COMB | 19 | -0.14 | 0.92 |  |
|  | Control | 14 | -0.09 | 0.95 |  |
| Visual Memory | AE | 25 | 0.17 | 1.05 | F(3,78) = 1.00, *p* =.398 |
|  | CCT | 23 | 0.06 | 0.98 |  |
|  | COMB | 19 | -0.34 | 1.17 |  |
|  | Control | 15 | 0.04 | 0.65 |  |
| Verbal Memory | AE | 25 | -0.04 | 0.71 | H(3) = 0.70, *p* = .874 |
|  | CCT | 23 | 0.16 | 0.79 |  |
|  | COMB | 19 | -0.04 | 1.08 |  |
|  | Control | 14 | -0.14 | 1.29 |  |
| Global Cognitive Function | AE | 23 | 0.01 | 0.73 | F(3,74) = 0.51, *p* = .675 |
|  | CCT | 23 | .011 | 0.56 |  |
|  | COMB | 19 | -0.13 | 0.57 |  |
|  | Control | 13 | -0.03 | 0.59 |  |
| AE = Aerobic exercise; CCT = Computerized Cognitive Training; COMB = Combined Training. | | | | | |

| **Table 2.2**  Group Comparison at Baseline: PA and CRF | | | | | |
| --- | --- | --- | --- | --- | --- |
| Variables | Groups | N | Mean | SD | ANOVA / H de Kruskall Wallis |
| CRF | AE | 19 | 25.25 | 10.16 | F(3,67) = 1.08, *p* = .362 |
|  | CCT | 20 | 26.11 | 12.50 |  |
|  | COMB | 17 | 27.34 | 8.75 |  |
|  | Control | 15 | 20.65 | 12.69 |  |
| S-PA | AE | 25 | 451.98 | 699.40 | H(3) = 2.92, *p* = .404 |
|  | CCT | 23 | 439.83 | 713.63 |  |
|  | COMB | 19 | 778.79 | 908.77 |  |
|  | Control | 15 | 366.80 | 618.17 |  |
| NS-PA | AE | 25 | 5595.73 | 3918.34 | H(3) = 7.96, *p* = .047* |
|  | CCT | 23 | 9113.74 | 7104.64 |  |
|  | COMB | 19 | 10295.68 | 6159.04 |  |
|  | Control | 15 | 7038.40 | 6628.45 |  |
| AE = Aerobic exercise; CCT = Computerized Cognitive Training; COMB = Combined Training; CRF = Cardiorespiratory Fitness; NS-PA = Non Sportive Physical Activity; S-PA = Sportive Physical Activity; Total-PA = Total Physical Activity.  **p* < 0.05 | | | | | |

| **Table 2.3**  Group Comparison at Baseline: Global Mean DC Strength estimated over the individual sFCN for the MOVIMENT PROJECTE and the NYU test-retest study | | | | | |
| --- | --- | --- | --- | --- | --- |
| Variables | Groups | N | Mean | SD | ANOVA / H de Kruskall Wallis |
| Global Mean DC Strength (MOVIMENT) | AE | 25 | 0.179 | 0.0052 | F(3,78) = 2.04, *p* = .125 |
|  | CCT | 23 | 0.176 | 0.0060 |  |
|  | COMB | 19 | 0.175 | 0.0050 |  |
|  | Control | 15 | 0.177 | 0.0060 |  |
| Global Mean DC Strength (NYU) | SCAN1 | 25 | 0.2060 | 0.002 | F(3,74) = 1.37, *p* = .259 |
|  | SCAN2 | 25 | 0.2070 | 0.002 |  |
|  | SCAN3 | 25 | 0.2060 | 0.002 |  |
| AE = Aerobic exercise; CCT = Computerized Cognitive Training; COMB = Combined Training; FA = Fractional Anisotropy; MD = Mean Diffusivity. | | | | | |

| **Table 3**  Comparison between baseline and follow-up intragroup: Global Mean DC Strength estimated over the individual sFCN | | | | |
| --- | --- | --- | --- | --- |
|  | **AE**  M(SD) Baseline - M(SD) Follow-up  t(d.f.); *p* Value | **CCT**  M(SD) Baseline - M(SD) Follow-up  t(d.f.); *p* Value | **COMB**  M(SD) Baseline - M(SD) Follow-up  t(d.f.); *p* Value | **Control**  (SD) Baseline - M(SD) Follow-up  t(d.f.); *p* Value |
| Global Mean DC Strength (MOVIMENT) | 0.179 (0.005) – 0.181 (0.008)  t(24)=2.88; *p*=.284 | 0.176 (0.006) – 0.178 (0.005)  t(22)=2.54; *p*=.304 | 0.175 (0.004) – 0.188 (0.005)  t(18)=2.12; *p*=.006 | 0.177 (0.006) – 0.178 (0.003)  t(14)=2.43; *p*=.189 |
| AE = Aerobic Exercise; CCT = Computerized Cognitive Training; COMB = Combined Training; M = Mean ; SD = Standard Deviation | | | | |
